# Investigating pleiotropy between depression and autoimmune diseases using the UK Biobank

**DOI:** 10.1101/2020.12.08.20242495

**Authors:** Kylie P Glanville, Jonathan R I Coleman, Paul F O’Reilly, James Galloway, Cathryn M Lewis

## Abstract

**Background:** Epidemiological studies have shown increased comorbidity between depression and autoimmune diseases. The mechanisms driving the comorbidity are poorly understood, and a highly powered investigation is needed to understand the relative importance of shared genetic influences. We investigated the evidence for pleiotropy from shared genetic risk alleles between these traits in the UK Biobank (UKB).

**Methods:** We defined autoimmune and depression cases using information from hospital episode statistics, self-reported conditions and medications, and mental health questionnaires. Pairwise comparisons of depression prevalence between autoimmune cases and controls, and vice-versa, were performed. Cross-trait polygenic risk score (PRS) analyses were performed to test for pleiotropy, i.e. testing whether PRS for depression could predict autoimmune disease status, and vice-versa.

**Results:** We identified 28k cases of autoimmune diseases (pooling across 14 traits) and 324k autoimmune controls, and 65k cases of depression and 232k depression controls. The prevalence of depression was significantly higher in autoimmune cases compared to controls, and vice-versa. PRS for myasthenia gravis and psoriasis were significantly higher in depression cases compared to controls (p < 5.2×10^−5^, *R*^*2*^ <= 0.04%). PRS for depression were significantly higher in inflammatory bowel disease, psoriasis, psoriatic arthritis, rheumatoid arthritis and type 1 diabetes cases compared to controls (p < 5.8×10^−5^, *R*^*2*^ range 0.06% to 0.27%), and lower in coeliac disease cases compared to controls (p < 5.4×10^−7^, *R*^*2*^ range 0.11% to 0.15%).

**Conclusions:** Consistent with the literature, depression was more common in individuals with autoimmune diseases compared to controls, and vice-versa, in the UKB. PRS showed some evidence for involvement of shared genetic factors, but the modest *R*^*2*^ values suggest that shared genetic architecture accounts for only a small proportion of the increased risk across traits.

## Introduction

There is evidence that individuals with a history of autoimmune disease are at greater risk for developing depression^1-4^, and that a history of depression increases risk for developing autoimmune diseases^5,6^. The mechanisms driving the bi-directional relationship are poorly understood, but one contributory factor may be that these diseases share biological pathways. We and others have shown that there is no strong evidence for the involvement of Human Leukocyte Antigen (HLA) alleles in risk for depression, suggesting that the Major Histocompatibility Complex (MHC) does not harbor shared risk for depression and autoimmune diseases^6-8^. However, genetic risk for autoimmune diseases occurs across the genome^9^, and pleiotropic effects outside the MHC may be involved in shared risk for depression and autoimmune diseases.

Few studies have investigated evidence for genome-wide pleiotropy between depression and autoimmune diseases. Euesden, et al.^10^ found no evidence for association between polygenic risk scores (PRS) for depression and risk for rheumatoid arthritis, or vice-versa. The Psychiatric Genomics Consortium (PGC) indicated no evidence for significant genetic correlations (rG) between depression and nine autoimmune diseases (after multiple testing correction across 221 traits in total); the strongest correlation observed was between depression and inflammatory bowel disease (rG = .07, uncorrected p = .01)^11^. Recently, Liu, et al.^6^ found no association between PRS for mental health disorders and risk for autoimmune diseases, and only a weak association between PRS for autoimmune diseases and risk for mental health disorders.

We extend previous work, leveraging the UK Biobank (UKB) to test for pleiotropy between depression and autoimmune diseases with PRS methodology. Given the challenge of reliably defining complex disease traits using large-scale data, we take two approaches to defining autoimmune diseases and depression. We classified liberally-defined cases, based on a single item endorsing diagnosis with an autoimmune disease, and strictly-defined cases, based on multiple items. We use this approach to identify individuals affected by any of fourteen autoimmune or autoinflammatory traits - collectively referred to as autoimmune diseases throughout. We take a similar approach to classifying depression by requiring a greater number of endorsements in strictly-defined cases than liberally-defined cases. Liberally-defined cases increase the sample size, while strictly-defined cases will reduce the rate of misclassification. We perform cross-trait PRS analyses, testing for association between PRS for autoimmune diseases and depression, and vice-versa. Motivated by the observation of sex-dependent genetic correlations between schizophrenia and autoimmune diseases^12^, and by higher prevalence in females of both depression and autoimmune diseases, we stratified PRS analyses by sex. Our study is one of the largest to explore pleiotropy between depression and autoimmune diseases and elucidates the contribution of shared genetic influences to the observed comorbidity.

## Methods

### Participants

The UKB is a prospective health study of 500,000 individuals in the United Kingdom. Participants were identified through NHS patient registers if they were aged 40-69 during the recruitment phase (2006-2010) and living in proximity to an assessment centre. Participants attended a baseline assessment and contributed health information via touchscreen questionnaires and verbal interviews^13^. Subsets of participants completed repeat assessments: instance 1) n = 20,335 between 2012-2013;instance 2) n = 42,961 (interview) and n = 48,340 (touchscreen) in 2014; and instance 3) n = 2,843 (interview) and n = 3,081 (touchscreen) in 2019. Participant data are linked to Hospital Episode Statistics (HES) containing information on episodes of inpatient care. Episodes are coded at admission using the International Classification of Diseases, 10^th^ Revision^14^ (ICD-10). Inpatients are assigned one primary code (reason for admission) and a variable number of secondary codes. Additional data are available for psychiatric phenotyping, including an online Mental Health Questionnaire (MHQ) completed by 157,366 participants in 2017^15^. The UKB received ethical approval from the North West - Haydock Research Ethics Committee (reference 16/NW/0274). Participants provided electronic signed consent at recruitment^13^.

### Autoimmune phenotyping

Guided by studies that investigated the epidemiological relationship between autoimmune diseases and depression^1^,^5^ we identified cases for fourteen autoimmune diseases: pernicious anemia (PA), autoimmune thyroid disease (ATD), type 1 diabetes (T1D), multiple sclerosis (MS), myasthenia gravis (MG), coeliac, inflammatory bowel disease (IBD; includes crohn’s disease and ulcerative colitis), psoriasis, ankylosing spondylitis (AS), polymyalgia rheumatica/giant cell arteritis (PR/GCA), psoriatic arthritis (PsA), rheumatoid arthritis (RA), sjögren syndrome (SS), and systemic lupus erythematosus (SLE).

Two sources of information were used to define autoimmune cases and controls. (1) HES: primary and secondary ICD-10 diagnoses recorded between April 1997 to October 2016 were identified from the UKB Data Portal Record Repository. (2) Verbal interview: participants’ responses at baseline or instance 1 or 2 to determine self-endorsed medical conditions (past and current) and self-endorsed prescription medications (current). ICD-10 codes, self-endorsed conditions and medications used to define each autoimmune disease are listed in the Supplementary Material.

We took two approaches to defining autoimmune cases (Figure 1). To increase sample size, we created ‘possible’ cases, comprising participants with an ICD-10 diagnosis or a self-endorsed condition. To increase validity, we used multiple observations to create ‘probable’ cases. Participants were coded as probable cases if at least two of ICD-10 diagnosis, self-endorsed condition or medication were observed. More than one ICD-10 diagnosis for the corresponding autoimmune disease was also sufficient. A set of autoimmune controls was defined from participants with no ICD-10 diagnoses, self-endorsed conditions or medications for all fourteen autoimmune diseases. A single set of controls was used for all autoimmune diseases, given the known comorbidity between them.

**Figure 1:**
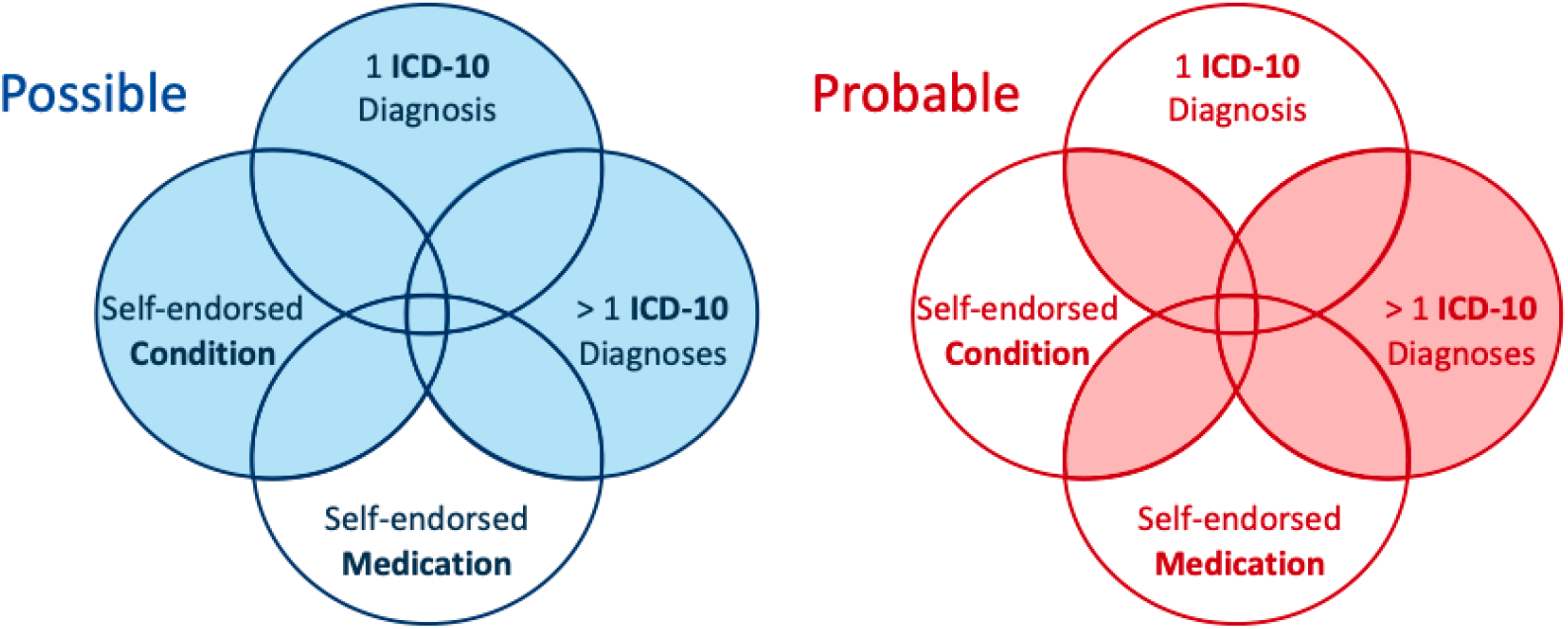
Autoimmune phenotyping approach. Cases are included in possible or probable if they fall within a shaded area. Autoimmune medication was used as a confirmatory, but not a primary source of information, because several medications are not disease-specific.

### Depression phenotyping

We created two depression case groups: strictly-defined cases termed ‘stringent depression’ and liberally-defined cases termed ‘any depression’. We have previously shown that SNP-based heritability increases with multiple endorsements of depression^16^. We therefore classified ‘stringent depression’ as participants endorsing at least three of the following depression measures: ICD-10 diagnoses (F32-F33.9); self-reported depression; self-reported antidepressant usage; single or recurrent depression (defined by Smith, et al.^17^ from responses to a questionnaire completed at baseline by 172,751 participants); or answered ‘yes’ to the questionnaire: “Have you ever seen a GP/psychiatrist for nerves, anxiety, tension or depression?”.

We classified ‘any depression’ as participants who endorsed two or more depression measures, or if they met criteria for lifetime depression in the Composite International Diagnostic Interview (CIDI) assessed in the MHQ^15^. We classify cases defined from CIDI alone as ‘any depression’ not as ‘stringent depression’ because we previously observed lower SNP-based heritability in this group (h^2^_SNP_ = 11%, SE = 0.008) compared to cases defined by three or more non-CIDI measures of depression (h^2^_SNP_ = 19%, SE = 0.018)^16^.

Depression cases were screened for schizophrenia and bipolar according to any indication: ICD-10 diagnoses (F20-29, F30-31.9, F34-39); self-endorsed conditions (schizophrenia, mania, bipolar disorder or manic depression) or self-endorsed antipsychotic usage reported at baseline or instance 1 or 2; Bipolar Type I (Mania) or Bipolar Type II (Hypomania) according to the criteria adopted by Smith, et al.^17^; or indications of psychosis endorsed in the MHQ. A single set of depression controls was defined from participants who did not meet the criteria for depression, schizophrenia or bipolar.

Derivation of depression, schizophrenia and bipolar indications can be found in Supplementary Materials from our previous publication^16^.

### Genetic quality control (QC)

The UKB performed preliminary QC on genotype data assayed for all participants^13^. Using genetic principal components (PCs) provided by the UKB, we performed 4-means clustering on the first two PCs to identify and retain individuals of European ancestry. QC was then performed using PLINK v1.9^18^ to remove: variants with missingness > 0.02 (before individual QC), individuals with missingness > 0.02, individuals whose self-reported sex was discordant from their genetic sex, variants with missingness > 0.02 (after individual QC), variants departing from Hardy-Weinberg Equilibrium (p < 10e-8), and variants with minor allele frequency (MAF) < 0.01. Relatedness kinship estimates provided by the UKB were used to identify pairs of related individuals (KING r^2^ > 0.044)^19^ and the GreedyRelated^20^ algorithm used to remove one individual from each pair, preferentially retaining individuals that survived QC. FlashPCA2^21^ was used to generate PCs for the sub-set of individuals of European ancestry surviving QC. PRS analyses were performed using genotype data.

### Statistical analyses

We summarised sociodemographic data taken at baseline assessment: age, sex, socio-economic status (SES), body mass index (BMI) and current smoking status. We tested for significant differences in sociodemographic variables between cases and controls using Welch Two Sample t-tests in R v3.6^22^. We tested for significant differences in: 1) the prevalence of depression in autoimmune cases compared to autoimmune controls, and 2) the prevalence of autoimmune diseases in depression cases compared to depression controls. These tests were performed for both probable/possible autoimmune cases and stringent/any depression, using 2-sample tests for equality of proportions in R v3.6^22^.

### Summary statistics for autoimmune diseases and depression

We searched PubMed and the NHGRI-EBI GWAS Catalog (https://www.ebi.ac.uk/gwas/downloads/summary-statistics) for the latest genome-wide association study (GWAS) with publicly-available summary statistics, using the name of the relevant trait (and “GWAS” or “genome-wide association study” on PubMed). We identified summary statistics for eight of the fourteen autoimmune diseases: coeliac^23^, IBD^24^, MS^25^, MG^26^, psoriasis^27^, PsA^28^, RA^29^, and SLE^30^ (Table 1). For MG, psoriasis and PsA, we contacted the authors of the primary GWASs directly to obtain access. Summary statistics from GWAS using the Immunochip were excluded as it does not provide genome-wide coverage. For Major Depressive Disorder (MDD), we used summary statistics from Wray, et al.^11^, excluding UKB.

**Table 1:**
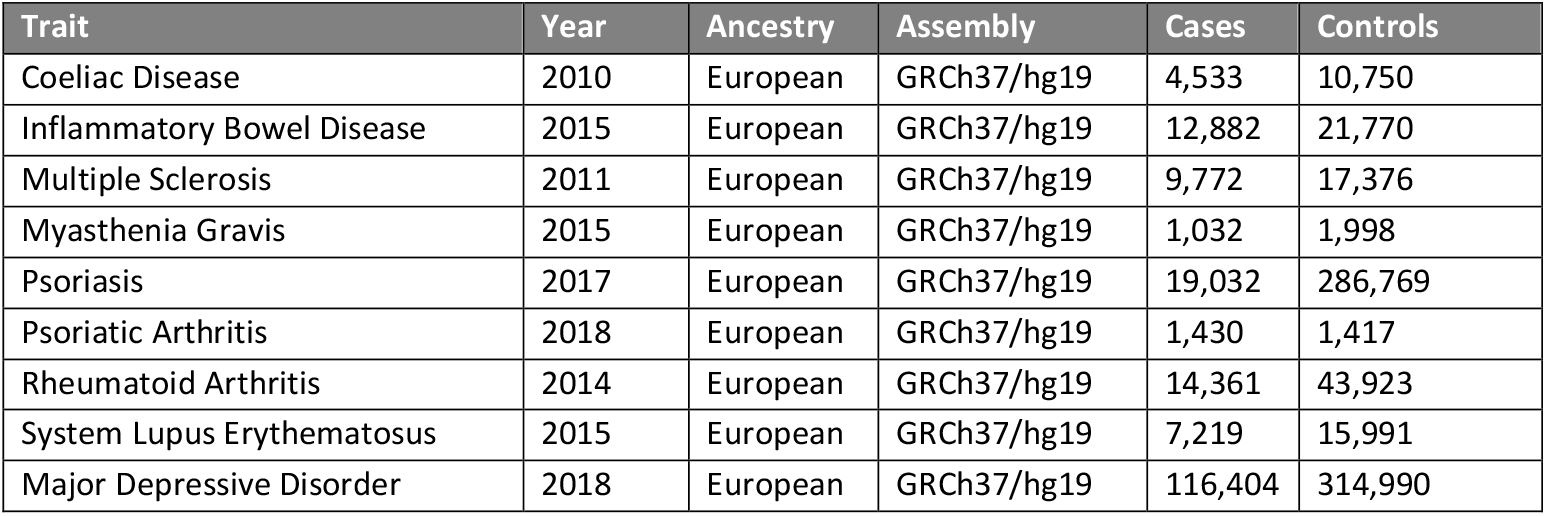
GWAS summary statistics used to generate polygenic risk scores.

### Polygenic risk score (PRS) analyses

PRS analyses were conducted using the PRSice-2 software^31^. QC was performed on summary statistics to remove variants within the MHC (28.8 to 33.7 Mb), and default clumping settings were applied in PRSice-2 to remove variants in linkage disequilibrium (r^2^ > 0.1) with the lead variant within a 250kb region.

To validate our phenotyping approach, we tested for association between PRS for eight autoimmune diseases and case-control status for the corresponding diseases (possible and probable cases), and between PRS for MDD and case-control status for depression (any and stringent).

To investigate pleiotropy between autoimmune diseases and depression we performed crosstrait analyses, testing for association between 1) PRS for eight autoimmune diseases and case-control status for depression (any and stringent cases); and 2) PRS for MDD and case-control status for fourteen autoimmune diseases (possible and probable cases). To test for sex-specific effects, we performed cross-trait analyses in males and females separately.

For each test, PRS constructed at eight p-value thresholds (*P*_T_; 0.001, 0.05, 0.1, 0.2, 0.3, 0.4, 0.5 and 1.0) were regressed on case-control status using logistic regression, adjusting for the following covariates: six PCs, genotyping batch, and assessment centre (*n*=128 variables). We report p-values at the optimal *P*_T_ for each test of association. To control for multiple testing across *P*_T_ (x8), and across tests of association (autoimmune PRS (x8) predicting any/stringent depression (x2) in men and women (x2), n=32; and MDD PRS predicting possible/probable (x2) autoimmune diseases (x14) in men and women (x2), n=56), a Bonferroni correction was applied to give a p-value threshold for significance of 7.1×10^−5^ (0.05/704 tests, 704 = 8*(32+56)). Where sex-specific associations were observed, sensitivity analyses were conducted to account for different sample sizes between sexes. We tested for interactions between sex and PRS (at the optimal *P*_T_ from sex-specific tests) in the full sample (Phenotype ∼ sex + PRS + sex*PRS + covariates). We report *R*^*2*^ estimates transformed to the liability scale using the following population prevalences for outcome traits: PA=0.1%^32^, ATD=2%^33^, T1D=0.3%^34^, MS=0.1%^25^, MG=0.02%^35^, coeliac=1%^23^, IBD=0.5%^24^, psoriasis=2%^36^, AS=0.55%^37^, PR/GCA= 0.85%^38^, PsA=0.5%^28^, RA=1%^39^, SS=0.7%^40^, SLE=0.1%^41^ and MDD=15%^11^.

AVENGEME^42^ was used to estimate power to detect cross-trait PRS associations, assuming varying degrees of genetic correlation (rG) between corresponding traits (rG 0.01 to 0.5). Power was estimated for cross-trait analyses where summary statistics for both traits were available (i.e. eight autoimmune disorders and MDD) so that SNP-based heritability (required for power calculations in AVENGEME) could be estimated using Linkage Disequilibrium Score Regression (LDSC v1.0.1)^43^ (Supplementary Figure 1). Power was estimated using PRS at the optimal *P*_T_ identified in cross-trait association tests, and liberally-defined sample sizes. Parameters used to estimate power are in Supplementary Tables 1 and 2.

LDSC v1.0.1^43^ was used to estimate rG between the UKB depression phenotypes (‘any’ and ‘stringent’) and autoimmune diseases with publicly-available summary statistics. To robustly apply LDSC, we limited the autoimmune diseases to those with sample sizes above 5k in the primary GWAS (coeliac^23^, IBD^24^, MS^25^, psoriasis^27^, RA^29^, and SLE^30^). To control for multiple testing across traits, a Bonferroni correction was applied to give a p-value threshold for significance of 4.1×10^−3^ in rG analyses (0.05/12 tests).

## Results

A total of 28,479 individuals were identified as possible cases across fourteen autoimmune diseases, and a sub-set of 16,824 (59.1%) met the stringent criteria for probable cases (refer Supplementary Material for representation of the overlap between criteria used to define cases). 65,075 individuals met the criteria for any depression, 14,625 of whom met the criteria for stringent depression. Sociodemographic characteristics for autoimmune and depression cases and controls are summarised in Table 2. Overall, autoimmune and depression case groups contained a higher proportion of females, had lower SES, higher smoking prevalence, and higher BMI than their respective control groups, (N = 324,074 autoimmune controls, N = 232,552 depression controls, all p-values < 5×10^−21^ in pairwise comparisons).

**Table 2:**
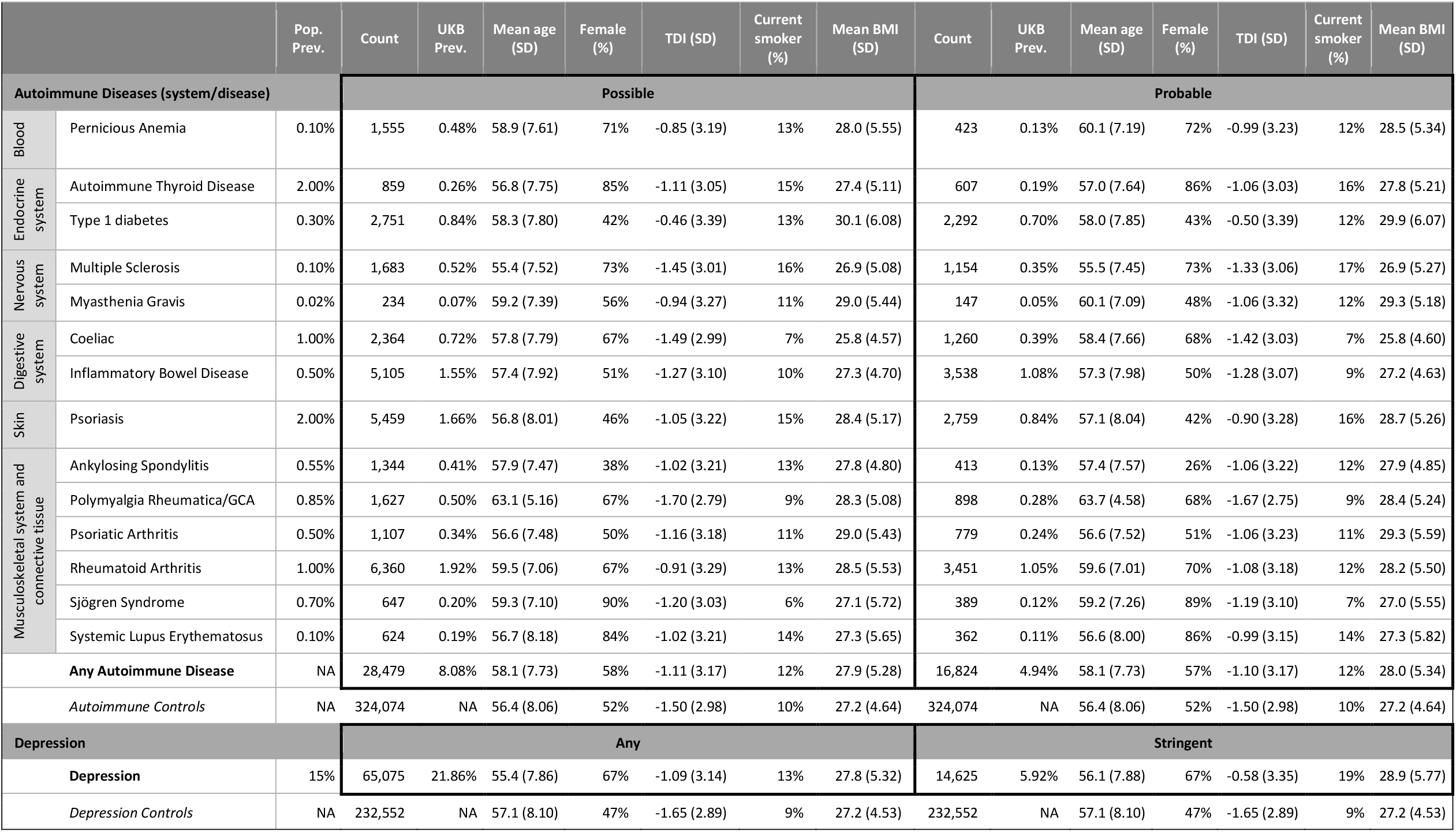
Sociodemographic information for autoimmune and depression cases and controls. Pop. Prev. = population prevalence estimate. UKB Prev. = prevalence of cases the UKB as a proportion of autoimmune/depression controls. TDI = Townsend Deprivation Index; negative scores indicate less deprivation. SD = standard deviation.

The prevalence of any depression was significantly higher in autoimmune cases compared to autoimmune controls (p = 6×10^−177^ for possible cases of any autoimmune disease versus controls, p = 2×10^−124^ for probable cases of any autoimmune disease versus controls). The prevalence of stringent depression was significantly higher in autoimmune cases compared to autoimmune controls (p = 3×10^−207^ for possible cases of any autoimmune disease versus controls, p = 6×10^−163^ for probable cases of any autoimmune disease versus controls) (Table 3).

**Table 3:** Prevalence of depression within autoimmune cases compared to autoimmune controls, stratified by possible/probable for autoimmune diseases, and any/stringent for depression cases. P-values from pairwise comparisons of depression prevalence in autoimmune cases compared to autoimmune controls are shown in brackets.

|  |  | Depression prevalence in autoimmune traits |  |  |  |
| --- | --- | --- | --- | --- | --- |
|  |  | Any depression prevalence |  | Stringent depression prevalence |  |
| Autoimmune Trait |  | Possible Autoimmune | Probable Autoimmune | Possible Autoimmune | Probable Autoimmune |
| **Autoimmune Controls** |  | **20.6%** |  | **5.1%** |  |
| **Any Autoimmune Disease** |  | **28.9% ($6 \times 10^{-177}$)** | **29.5% ($2 \times 10^{-124}$)** | **10.8% ($3 \times 10^{-207}$)** | **11.5% ($6 \times 10^{-163}$)** |
| Blood | Pernicious Anemia | 35.8% ($4 \times 10^{-36}$) | 33.7% ($4 \times 10^{-8}$) | 15.2% ($1 \times 10^{-39}$) | 16.6% ($4 \times 10^{-15}$) |
| Endocrine system | Autoimmune Thyroid Disease | 35.1% ($6 \times 10^{-19}$) | 34.7% ($6 \times 10^{-13}$) | 13.7% ($1 \times 10^{-16}$) | 13.4% ($3 \times 10^{-11}$) |
| | Type 1 diabetes | 31.1% ($3 \times 10^{-31}$) | 31.3% ($1 \times 10^{-27}$) | 14.1% ($5 \times 10^{-59}$) | 13.5% ($5 \times 10^{-43}$) |
| Nervous System | Multiple Sclerosis | 39.3% ($6 \times 10^{-56}$) | 42.5% ($3 \times 10^{-51}$) | 16.9% ($5 \times 10^{-54}$) | 20.0% ($8 \times 10^{-57}$) |
| | Myasthenia Gravis | 33.3% ($5 \times 10^{-5}$) | 29.2% ($3 \times 10^{-2}$) | 14.1% ($5 \times 10^{-6}$) | 14.8% ($4 \times 10^{-5}$) |
| Digestive system | Coeliac | 27.3% ($3 \times 10^{-12}$) | 29.8% ($2 \times 10^{-12}$) | 8.4% ($4 \times 10^{-8}$) | 9.5% ($7 \times 10^{-8}$) |
| | Inflammatory Bowel Disease | 24.9% ($6 \times 10^{-11}$) | 24.9% ($3 \times 10^{-8}$) | 8.9% ($8 \times 10^{-22}$) | 9.1% ($2 \times 10^{-17}$) |
| Skin | Psoriasis | 26.8% ($2 \times 10^{-22}$) | 28.3% ($1 \times 10^{-17}$) | 8.7% ($4 \times 10^{-20}$) | 9.9% ($4 \times 10^{-18}$) |
| Musculoskeletal system and connective tissue | Ankylosing Spondylitis | 26.9% ($5 \times 10^{-7}$) | 28.4% ($5 \times 10^{-4}$) | 9.6% ($7 \times 10^{-9}$) | 10.5% ($9 \times 10^{-5}$) |
| | Polymyalgia Rheumatica/GCA | 29.3% ($2 \times 10^{-13}$) | 27.8% ($5 \times 10^{-6}$) | 10.8% ($3 \times 10^{-15}$) | 10.6% ($1 \times 10^{-8}$) |
| | Psoriatic Arthritis | 31.8% ($3 \times 10^{-15}$) | 34.3% ($5 \times 10^{-16}$) | 10.2% ($1 \times 10^{-8}$) | 13.0% ($2 \times 10^{-13}$) |
| | Rheumatoid Arthritis | 30.6% ($5 \times 10^{-62}$) | 29.4% ($6 \times 10^{-28}$) | 12.7% ($3 \times 10^{-92}$) | 12.1% ($1 \times 10^{-45}$) |
| | Sjögren Syndrome | 40.7% ($4 \times 10^{-25}$) | 41.9% ($1 \times 10^{-17}$) | 18.9% ($2 \times 10^{-28}$) | 18.9% ($7 \times 10^{-18}$) |
| | Systemic Lupus Erythematosus | 40.2% ($2 \times 10^{-25}$) | 44.5% ($2 \times 10^{-22}$) | 18.8% ($6 \times 10^{-30}$) | 22.2% ($8 \times 10^{-27}$) |

The prevalence of possible cases of any autoimmune disease was significantly higher in depression cases compared to depression controls (p = 6×10^−177^ for any depression versus controls, p = 3×10^−207^ for stringent depression versus controls). The prevalence of probable cases of any autoimmune disease was significantly higher in depression cases compared to depression controls (p = 2×10^−124^ for any depression versus controls, p = 6×10^−163^ for stringent depression versus controls) (Table 4).

**Table 4:** Prevalence of autoimmune diseases within depression cases compared to depression controls, stratified by possible/probable for autoimmune diseases, and stringent/any for depression cases. P-values from pairwise comparisons of autoimmune prevalence in depression cases compared to depression controls are shown in brackets.

|  |  | Autoimmune prevalence in depression |  |  |  |  |  |
| --- | --- | --- | --- | --- | --- | --- | --- |
|  |  | Possible autoimmune prevalence |  |  | Probable autoimmune prevalence |  |  |
| Autoimmune Trait |  | Any Depression | Stringent Depression | Depression Controls | Any Depression | Stringent Depression | Depression Controls |
| **Any Autoimmune Disease** | | **10.48%** ($6 \times 10^{-177}$) | **14.27%** ($3 \times 10^{-207}$) | **6.94%** | **6.59%** ($2 \times 10^{-124}$) | **9.48%** ($6 \times 10^{-163}$) | **4.19%** |
| Blood | Pernicious Anemia | 0.77% ($4 \times 10^{-36}$) | 1.17% ($1 \times 10^{-39}$) | 0.36% | 0.19% ($4 \times 10^{-8}$) | 0.35% ($4 \times 10^{-15}$) | 0.10% |
| Endocrine system | Autoimmune Thyroid Disease | 0.41% ($6 \times 10^{-19}$) | 0.58% ($1 \times 10^{-16}$) | 0.20% | 0.28% ($6 \times 10^{-13}$) | 0.39% ($3 \times 10^{-11}$) | 0.14% |
| | Type 1 diabetes | 1.20% ($3 \times 10^{-31}$) | 2.07% ($5 \times 10^{-59}$) | 0.69% | 1.02% ($1 \times 10^{-27}$) | 1.65% ($5 \times 10^{-43}$) | 0.58% |
| Nervous System | Multiple Sclerosis | 0.87% ($6 \times 10^{-56}$) | 1.31% ($5 \times 10^{-54}$) | 0.35% | 0.63% ($3 \times 10^{-51}$) | 1.02% ($8 \times 10^{-57}$) | 0.22% |
| | Myasthenia Gravis | 0.11% ($5 \times 10^{-5}$) | 0.17% ($5 \times 10^{-6}$) | 0.05% | 0.06% ($3 \times 10^{-2}$) | 0.12% ($4 \times 10^{-5}$) | 0.04% |
| Digestive system | Coeliac | 0.92% ($3 \times 10^{-12}$) | 1.08% ($4 \times 10^{-8}$) | 0.64% | 0.54% ($2 \times 10^{-12}$) | 0.64% ($7 \times 10^{-8}$) | 0.33% |
| | Inflammatory Bowel Disease | 1.82% ($6 \times 10^{-11}$) | 2.56% ($8 \times 10^{-22}$) | 1.43% | 1.28% ($3 \times 10^{-8}$) | 1.85% ($2 \times 10^{-17}$) | 1.00% |
| Skin | Psoriasis | 2.05% ($2 \times 10^{-22}$) | 2.55% ($4 \times 10^{-20}$) | 1.46% | 1.10% ($1 \times 10^{-17}$) | 1.47% ($4 \times 10^{-18}$) | 0.73% |
| Musculoskeletal system and connective tissue | Ankylosing Spondylitis | 0.52% ($5 \times 10^{-7}$) | 0.72% ($7 \times 10^{-9}$) | 0.37% | 0.17% ($5 \times 10^{-4}$) | 0.25% ($9 \times 10^{-5}$) | 0.11% |
| | Polymyalgia Rheumatica/GCA | 0.66% ($2 \times 10^{-13}$) | 0.93% ($3 \times 10^{-15}$) | 0.42% | 0.35% ($5 \times 10^{-6}$) | 0.51% ($1 \times 10^{-8}$) | 0.23% |
| | Psoriatic Arthritis | 0.50% ($3 \times 10^{-15}$) | 0.58% ($1 \times 10^{-8}$) | 0.28% | 0.38% ($5 \times 10^{-16}$) | 0.51% ($2 \times 10^{-13}$) | 0.19% |
| | Rheumatoid Arthritis | 2.66% ($5 \times 10^{-62}$) | 4.14% ($3 \times 10^{-92}$) | 1.59% | 1.42% ($6 \times 10^{-28}$) | 2.23% ($1 \times 10^{-45}$) | 0.89% |
| | Sjögren Syndrome | 0.34% ($4 \times 10^{-25}$) | 0.55% ($2 \times 10^{-28}$) | 0.13% | 0.21% ($1 \times 10^{-17}$) | 0.33% ($7 \times 10^{-18}$) | 0.08% |
| | Systemic Lupus Erythematosus | 0.36% ($2 \times 10^{-25}$) | 0.59% ($6 \times 10^{-30}$) | 0.14% | 0.23% ($2 \times 10^{-22}$) | 0.39% ($8 \times 10^{-27}$) | 0.08% |

Testing for same-trait PRS associations, PRS for MDD were significantly associated with any depression case-status (p < 5×10^−324^, *R*^*2*^ = 1.48%,) and stringent depression case-status (p = 2×10^−228^, *R*^*2*^ = 2.23%). PRS for autoimmune diseases were significantly associated with both possible and probable case-control status for the corresponding diseases (Figure 2). The variance in liability, *R*^*2*^, explained by PRS was higher in strictly-defined compared to liberally-defined phenotypes. Most results were highly significant (p < 6×10^−29^), except myasthenia gravis (p < 7×10^−3^), which has the smallest sample size of 234 cases, and psoriatic arthritis (p < 3 x 10^−6^) where the discovery GWAS has only 1430 cases.

**Figure 2:**
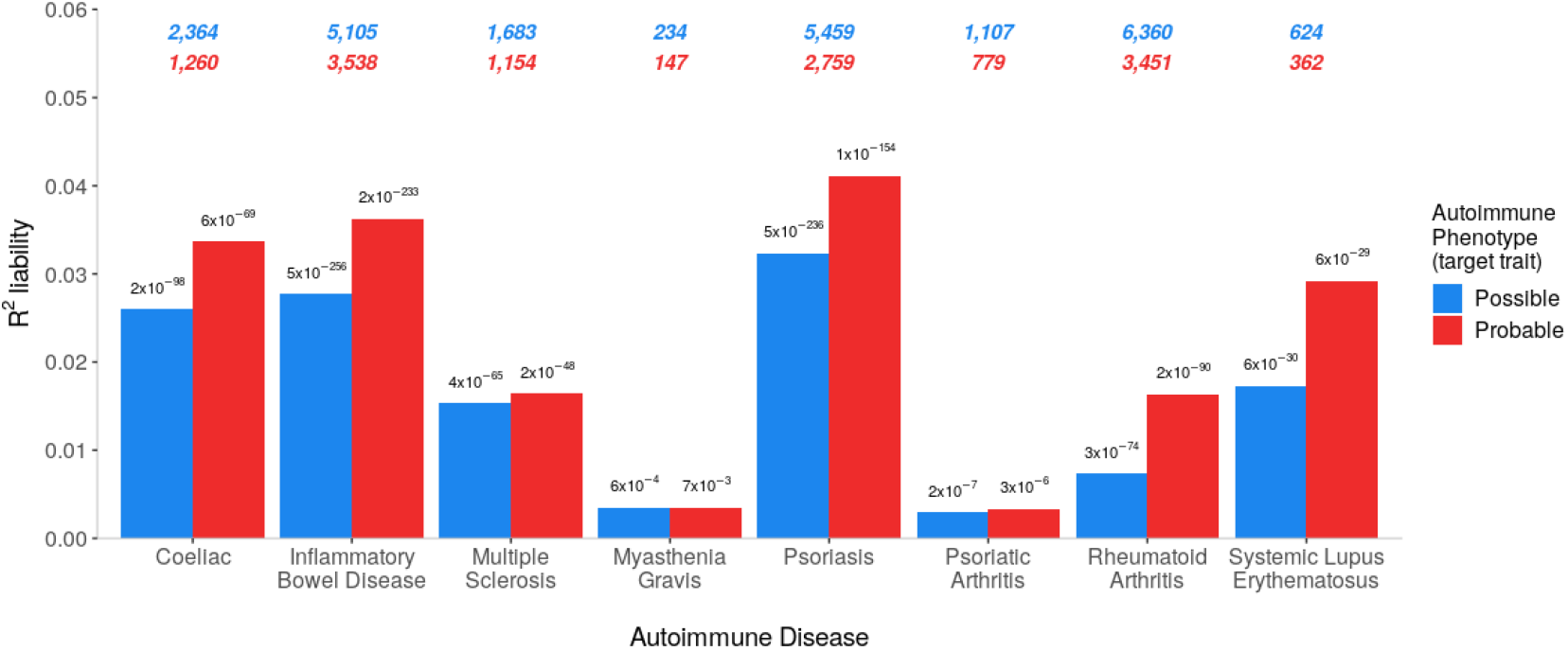
Variances in autoimmune liability explained by PRS for the corresponding autoimmune diseases. The number of cases are shown at the top of the plot (possible = blue, probable = red). P-values are shown atop each bar.

Power analyses showed that, in the prediction of any depression from autoimmune PRS, there was 80% power to detect associations assuming modest levels of underlying genetic correlation (rG); rG <0.05 for coeliac, MS, psoriasis, and SLE; rG <0.1 for IBD, PsA and RA; and rG < 0.17 for MG (Supplementary Figure 2). In the prediction of possible autoimmune diseases from depression PRS, there was 80% power to detect associations assuming rG < 0.05 for coeliac and IBD; rG < 0.1 for psoriasis and RA; and rG < 0.15 for PsA and SLE. There were two exceptions; MS and MG, where the underlying rG would need to approach ∼0.3 to achieve 80% power (Supplementary Figure 3).

In the prediction of depression from autoimmune PRS (Figure 3), PRS for myasthenia gravis were significantly associated with case-status for any depression (p = 5.2×10^−5^, *R*^*2*^ = 0.01%) and stringent depression (p = 1.6×10^−5^, *R*^*2*^ = 0.04%). PRS for psoriasis were significantly associated with case-status for any depression (p = 8.7×10^−6^, *R*^*2*^ = 0.01%). No other autoimmune disease PRS predicted depression case-control status, and no sex-specific analyses met the Bonferroni-corrected threshold. The *R*^*2*^ values for variance explained in depression by autoimmune PRS are all very low, at <0.1%, and substantially lower than the *R*^*2*^ for autoimmune diseases (Figure 2).

**Figure 3:**
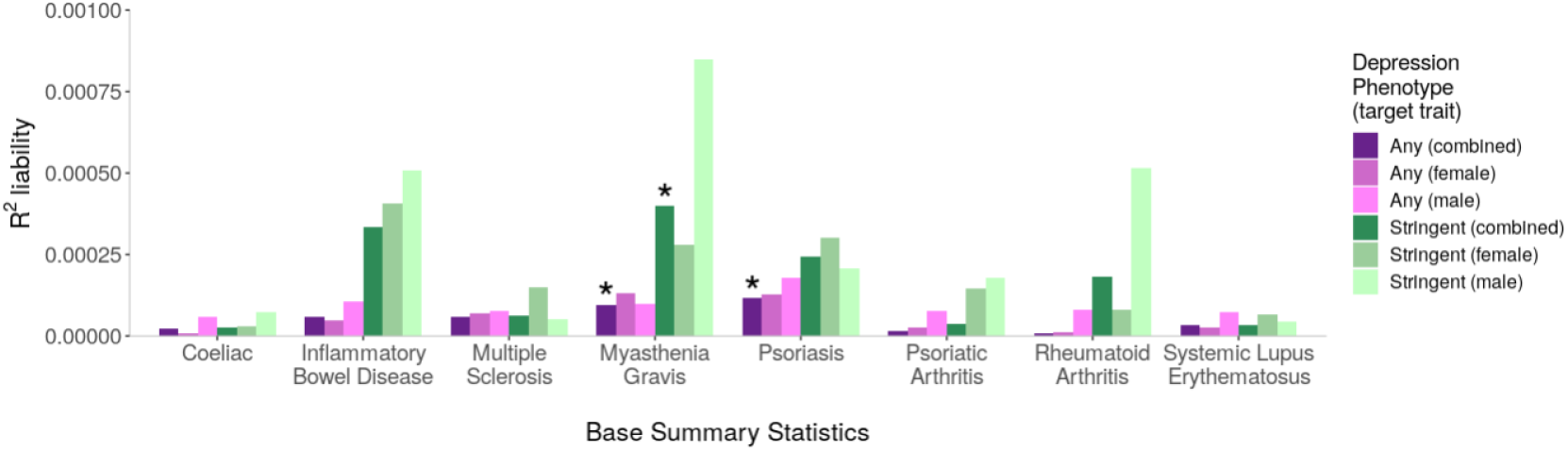
Variances in depression liability explained by PRS for autoimmune diseases (x-axis). Asterisks denote associations with p-values < 7.1×10^−5^, meeting Bonferroni correction. Number of cases for depression phenotypes: Any (combined) = 65,075; Any (female) = 43,413; Any (male) = 21,662; Stringent (combined) = 14,625; Stringent (female) = 9,738; Stringent (male) = 4,887.

In the prediction of autoimmune diseases from depression PRS, genetic liability for MDD was significantly associated with six autoimmune diseases: coeliac, inflammatory bowel disease, psoriasis, psoriatic arthritis, rheumatoid arthritis, and type 1 diabetes (all p-values < 5.8×10^−5^, *R*^*2*^ range between 0.06% and 0.27%) (Figure 4). For three, the association with MDD was observed in probable and possible cases (psoriasis, rheumatoid arthritis and type 1 diabetes). For coeliac and inflammatory bowel disease, the association was only in possible cases. For psoriatic arthritis, the association was only in probable cases. For all significant associations, higher PRS increased risk for the outcome phenotype, except for coeliac, where higher MDD PRS was associated with reduced risk (p = 6×10^−8^, *R*^*2*^ = 0.17%, beta = −0.11, SE = 0.02, in the combined sample).

**Figure 4:**
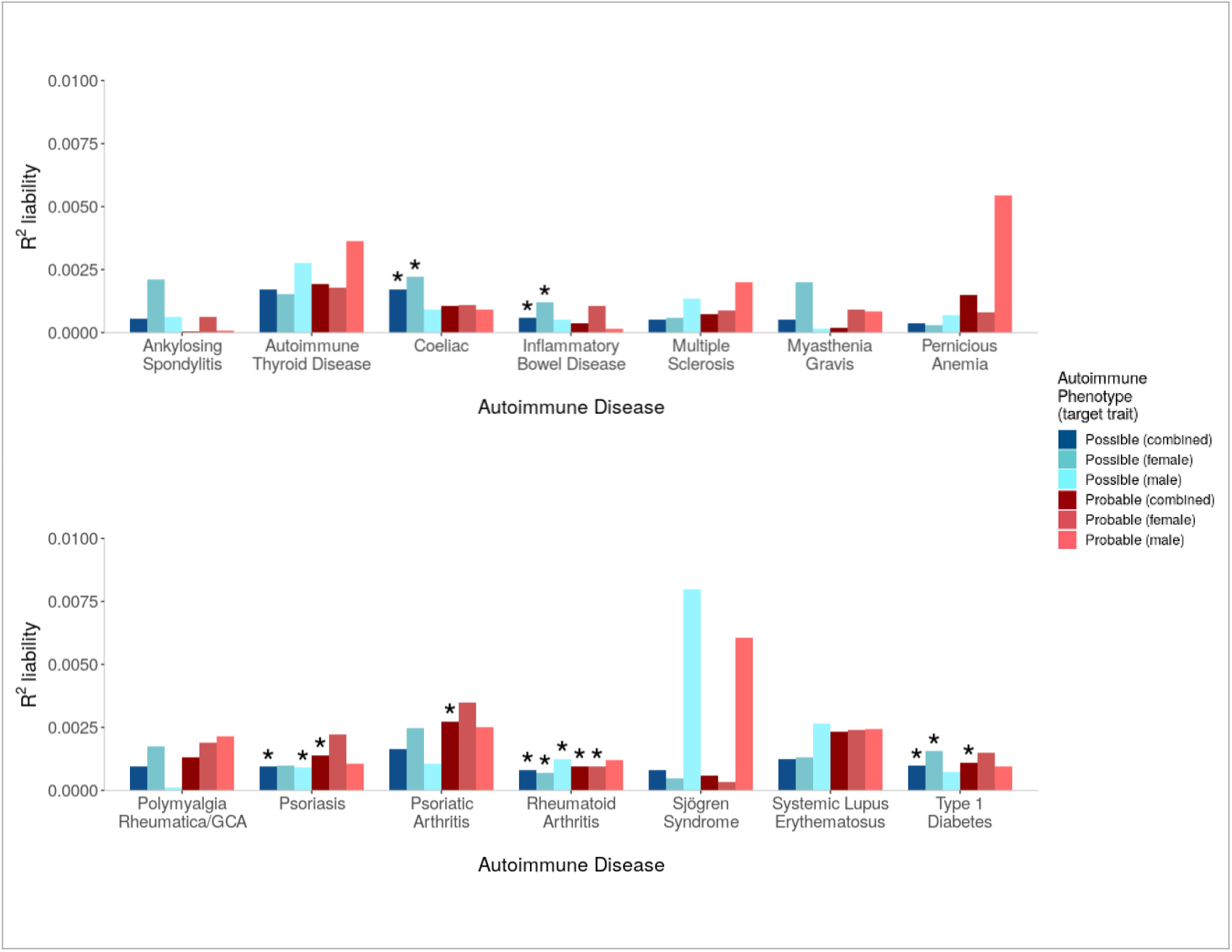
Variances in autoimmune liability (x-axes) explained by PRS for MDD. Asterisks denote associations with p-values < 7.1×10^−5^, meeting Bonferroni correction. Number of cases for the autoimmune diseases are given in Table 2.

In the prediction of autoimmune diseases from depression PRS, sex-specific associations were observed, primarily in female autoimmune cases (coeliac, inflammatory bowel disease, type 1 diabetes and rheumatoid arthritis, all p < 4.5×10^−5^). Association in males was observed in psoriasis (possible cases, p = 5.8×10^−5^), and in rheumatoid arthritis (possible cases, p = 1.6×10^−5^). The most consistent results were observed in rheumatoid arthritis, where the sample size was largest, with five of the six analyses reaching Bonferroni threshold (all p < 4.5×10^−5^, *R*^*2*^ range between 0.07% and 0.1%). However, there was no evidence for a significant interaction between sex and PRS in the combined samples of men and women (all p > 0.02), indicating that sex-specific associations were generally influenced by sample size.

Full results of each test are shown in Supplementary Tables 3 to 6 and Supplementary Figures 4 to 7.

Significant genetic correlations (rG) were observed between inflammatory bowel disease and any depression (rG = 0.11, 95% CI = 0.03 - 0.18, p = 3.8×10^−3^) and stringent depression (rG = 0.16, 95% CI = 0.07 - 0.24, p = 3.0×10^−4^); and between psoriasis and stringent depression (rG = 0.16, 95% CI = 0.06 - 0.26, p = 1.1×10^−3^). No other traits met the Bonferroni-corrected threshold for significance in rG analyses (Supplementary Table 7).

## Discussion

Motivated by epidemiological findings of a bi-directional relationship between depression and autoimmune diseases, we tested for evidence of pleiotropy between traits, adopting both liberal and strict phenotyping to define cases in the UKB. We showed modest association of PRS from autoimmune diseases with MDD, and slightly stronger associations of MDD PRS with autoimmune diseases. These observations suggest only a minor component of observed comorbidity is due to shared genetics between depression and autoimmune diseases.

We made three key observations: 1) Phenotypic variance explained by PRS for corresponding traits was higher in strictly-defined than liberally-defined cases, indicating more rigorous phenotyping improved the validity of autoimmune and depression cases; 2) The phenotypic overlap between depression and autoimmune diseases was consistent with the literature – depression was more common in individuals with autoimmune diseases, and vice-versa; 3) Cross-trait PRS analyses identified significant associations between depression and some autoimmune diseases, but with effect sizes indicating the existence of a shared biological component of modest effect on the observed comorbidity.

Our phenotyping approach used both strictly-defined and liberally-defined cases, integrating the multiple sources of UKB data. PRS for eight autoimmune diseases predicted case-control status, increasing confidence in the robustness of case definition. The phenotypic variance explained was higher in strictly-defined cases, potentially reflecting greater specificity; identifying individuals with multiple endorsements for a disease reduces the probability of misclassifying controls as cases. Conversely, the criteria for liberally-defined cases increases sample size, but may induce misclassification of controls as cases.

For each of the autoimmune diseases considered, cases had higher frequencies of depression than controls, recapitulating the effect observed in epidemiological studies. Similarly, the prevalence of each autoimmune disease was significantly higher in depression cases compared to controls. Prevalence estimates reported here are cross-sectional, and we lack information on the temporal relationship between traits.

Cross-trait PRS analyses identified significant associations, although observed effect sizes were small, ranging between *R*^*2*^ = 0.01% and 0.27%. Compared with the substantially higher phenotypic variance explained by PRS for autoimmune diseases in corresponding traits, the small effect sizes observed in cross-trait PRS analyses provide a useful contrast, indicating only a small contribution of shared genetic influences in the observed comorbidities. However, this was not universally true – MDD PRS captured nearly the same amount of variance in probable psoriatic arthritis (0.27%) as the PRS for psoriatic arthritis (0.29%). For all significant associations, higher PRS increased risk for the outcome phenotype. Interestingly, there was one exception, where higher MDD PRS was associated with reduced risk for coeliac disease. This is intriguing given the positive phenotypic correlation between depression and coeliac disease and may warrant further investigation.

For three traits, we observed significant associations in liberally-defined, but not strictly-defined cases (psoriasis PRS was associated with any depression, MDD PRS was associated with possible coeliac and inflammatory bowel disease). In contrast, MDD PRS was associated with probable, but not possible, psoriatic arthritis, suggesting misclassification in possible cases. Misclassification bias may vary across diseases; some autoimmune diseases may be more prone to misclassification with other autoimmune diseases, whilst other diagnoses may misclassify with non-inflammatory conditions. For example, osteoarthritis (non-inflammatory) may misclassify as psoriatic arthritis in the absence of multiple-item endorsement to increase diagnosis validity.

Cross-trait PRS analyses identified some sex-dependent associations. MDD PRS were associated with psoriasis in males, and MDD PRS were associated with coeliac, inflammatory bowel disease and type 1 diabetes in females. However, sensitivity analyses revealed no evidence for significant interactions between PRS and sex in the combined sample, indicating that sex-dependent associations were generally driven by different sample sizes in sex-stratified analyses. Rheumatoid arthritis was the most common autoimmune disease and showed the most consistency in cross-trait associations; MDD PRS were significantly associated with rheumatoid arthritis in all case groups, except probable males. This is in contrast with Euesden, et al.^10^, found no evidence for association between PRS for depression and risk for rheumatoid arthritis, but in a substantially smaller sample of 226 cases. Liu, et al.^6^ also found no evidence for association between a composite mental health disorder PRS and risk for autoimmune diseases, but also in a smaller sample of 1,383 individuals with any of seven autoimmune diseases. A composite PRS for autoimmune diseases did show weak association with case-control status in a sample of 43,902 individuals with any of six mental health disorders in the Liu, et al.^6^ study. This highlights the importance of sample size, and our study benefits from the scale of the UKB, where power calculations indicated our investigation was able to detect modest pleiotropic effects.

In contrast to the small, but significant, cross-trait PRS associations observed between depression and several autoimmune diseases, we only observed significant genetic correlations between depression and two autoimmune diseases: inflammatory bowel disease and psoriasis. The PRS methodology, which exploits the use of individual-level data, may have increased power to detect weak genetic effects compared to LDSC, which uses only summary statistics.

The weak genetic contribution suggests that another mechanism may be driving or contributing to the bi-directional relationship between autoimmune diseases and depression^44^. Inflammatory factors underlying some cases of depression could provide a common biological pathogenesis with autoimmune diseases. Lynall, et al.^45^ observed increased immune cell counts in depression cases compared to controls, and identified a sub-group of cases with elevated inflammatory markers who presented with more severe depression than uninflamed cases. Environmental risk factors such as BMI and childhood maltreatment increase risk of both depression and autoimmune diseases and would contribute to the bi-directional effect^46,47^. Similarly, some treatments for depression (antidepressants) and autoimmune diseases (steroids) are obesogenic and may increase comorbidity. Diagnosis with autoimmune disease increases risk of depression due to psychological factors in adjusting to a chronic disorder and changes in behaviour such as reduced exercise. Health related behaviours that are elevated in depression (smoking, poor diet and reduced physical activity) increase risk for autoimmune diseases. These mechanisms may not be independent of joint genetic contributors. For example, shared inflammatory mechanisms would lead to horizontal pleiotropy, where genetic variants directly affect both disorders, and vertical pleiotropy can arise through environmental risk factors where genetic variation influences one trait through mediation on another trait. The mechanisms underpinning the observed cross-trait PRS associations may warrant further investigation, potentially using Mendelian Randomization to investigate whether MDD risk alleles have a causal effect on autoimmune diseases, and vice versa. It is also interesting to speculate that associations could be driven by ‘phenotypic hitchhiking’, in which a GWAS for one trait (e.g. MDD) ascertains patients with comorbid diseases (e.g. autoimmune), potentially inducing cross-trait correlations. Disentangling pleiotropy from ‘phenotypic hitchhiking’ may warrant further investigation.

## Limitations

A healthy volunteer bias has been observed in the UKB^48^, and is a noted limitation of the study. However, it has been proposed that this bias may attenuate, but not invalidate, exposure-outcome relationships^49^. A further limitation of the ability to extrapolate our results is the lack of representation in individuals of diverse ancestries. The literature has demonstrated attenuation in PRS analyses where training and target samples are drawn from different ancestral populations^50^, highlighting the need to perform GWAS in diverse ancestries. This limitation may have broader implications than would otherwise be the case for some conditions, such as SLE, which disproportionately affect individuals of African and Asian ancestry.

Although every effort has been made to address the potential for misclassification bias through the criteria for multiple-item endorsements in strictly-defined cases, the approach remains imperfect. For example, limited sample size led to us combine Thyroiditis and Grave’s disease, which have opposing thyroid function, under the broader classification of autoimmune thyroid disease.

Despite the scale of the UKB, power calculations showed that for some rare autoimmune diseases, larger samples would be required to reject the presence of a weak genetic correlation with depression. We also observed low SNP-based heritability using published summary statistics for multiple sclerosis, which reduced power to detect pleiotropic effects. The Bonferroni correction applied to cross-trait PRS analyses was conservative since the eight PRS p-value thresholds included in each test of association are correlated, although it is difficult to determine the appropriate correction and we chose to be strict rather than liberal.

## Conclusions

We identified cases and controls for depression and fourteen autoimmune diseases in the UKB, using both strict and liberal phenotyping. PRS analyses indicated that strict phenotyping improved the validity of cases, demonstrating that multiple UKB variables can be leveraged to increase specificity. Consistent with the literature, we found that depression was enriched in autoimmune cases, and vice-versa. Despite having power to detect subtle pleiotropic effects, we found little evidence that shared genetic factors have a meaningful influence on the observed co-occurrence of depression and autoimmune diseases in the UK Biobank. The limited shared genetic component will make only a modest contribution to the bidirectional disease risks, and shared environmental factors, including health-related characteristics and stressful life events, may be important. Future studies leveraging phenotypic, genetic, diagnostic, treatment and environmental risk factors may be necessary to unpick the mechanisms contributing to shared risks for autoimmune diseases and depression. In particular, future research should consider the psychological impacts of autoimmune disease while remaining cognizant of the need to consider and treat the two diseases in parallel.

## Data Availability

Available from UK Biobank subject to standard procedures (www.ukbiobank.ac.uk). The full GWAS summary statistics for the 23andMe discovery data set will be made available through 23andMe to qualified researchers under an agreement with 23andMe that protects the privacy of the 23andMe participants. Please visit https://research.23andme.com/collaborate/#publication for more information and to apply to access the data.

https://research.23andme.com/collaborate/#publication

## Acknowledgements

We thank participants and scientists involved in making the UK Biobank resource available (http://www.ukbiobank.ac.uk/). The UKB received ethical approval from the North West – Haydock Research Ethics Committee (reference 16/NW/0274). This study was conducted under application number 18177. We thank the research participants and employees of 23andMe for making this work possible. The MDD GWAS summary statistics results from 23andMe were available through a Data Transfer Agreement between 23andMe, Inc., and King’s College, London. Only summary statistics were shared with no individual level data. 23andMe participants provided informed consent and participated in the research online. The 23andMe protocol was approved by an external Association for the Accreditation of Human Research Protection Programs accredited Institutional Review Board, Ethical and Independent Review Services. Participants were included in the analysis on the basis of consent status as checked at the time data analyses were initiated. Statistical analyses were carried out on the King’s Health Partners High Performance Compute Cluster funded with capital equipment grants from the GSTT Charity (TR130505) and Maudsley Charity (980). We thank Nick Dand, Satveer Mahil and Catherine Smith of King’s College London for their contribution in identifying medications used in the treatment of Psoriasis in the UKB.

## Data Code and Availability

Available from UK Biobank subject to standard procedures (www.ukbiobank.ac.uk).The full GWAS summary statistics for the 23andMe discovery data set will be made available through 23andMe to qualified researchers under an agreement with 23andMe that protects the privacy of the 23andMe participants. Please visit https://research.23andme.com/collaborate/#publication for more information and to apply to access the data.

The code used during this study are available at GitHub: https://github.com/kglanville/pleiotropy_autoimmune_depression_ukb.

## Author contributions

Conceptualisation and study design: KPG, CML, JG, PFO. Analysis and manuscript: KPG. Analytical consultation and interpretation: CML, JG, PFO, JRIC. UKB data curation and management: JRIC, KPG. Genetic data preparation: JRIC, KPG. Project supervisors: CML, JG, PFO. All authors critically edited the paper.

## References

1. Benros ME, Waltoft BL, Nordentoft M, Ostergaard SD, Eaton WW, Krogh J, et al. Autoimmune Diseases and Severe Infections as Risk Factors for Mood Disorders A Nationwide Study. JAMA Psychiatry. American Medical Association; 2013 Aug;70(8):812–20.

2. Anderson RJ, Freedberg KA, Clouse RE, Lustman PJ. The Prevalence of Comorbid Depression in Adults With Diabetes. Diabetes Care. 2001 Jun;24(6):1069–78.

3. Patten SB, Beck CA, Williams JVA, Barbui C, Metz LM. Major depression in multiple sclerosis: A population-based perspective. Neurology. 61st ed. 2003 Dec;:1524–7.

4. Kurina L, Goldacre M, Yeates D, Gill L. Depression and anxiety in people with inflammatory bowel disease. Journal of Epidemiology and Community Health. BMJ Publishing Group; 2001 Oct 1;55(10):716–20.

5. Andersson NW, Gustafsson LN, Okkels N, Taha F, Cole SW, Munk-Jorgensen P, et al. Depression and the risk of autoimmune disease: a nationally representative, prospective longitudinal study. Psychol Med. Cambridge University Press; 2015 Dec;45(16):3559– 69.

6. Liu X, Nudel R, Thompson WK, Appadurai V, Schork AJ, Buil A, et al. Genetic factors underlying the bidirectional relationship between autoimmune and mental disorders – findings from a Danish population-based study. Brain, Behavior, and Immunity. Academic Press; 2020 Jun 11.

7. Glanville KP, Coleman JRI, Hanscombe KB, Euesden J, Choi SW, Purves KL, et al. Classical Human Leukocyte Antigen Alleles and C4 Haplotypes Are Not Significantly Associated With Depression. Biological Psychiatry. Elsevier; 2020 Mar 1;87(5):419–30.

8. Nudel R, Benros ME, Krebs MD, Allesøe RL, Lemvigh CK, Bybjerg-Grauholm J, et al. Immunity and mental illness: findings from a Danish population-based immunogenetic study of seven psychiatric and neurodevelopmental disorders. European Journal of Human Genetics. Nature Publishing Group; 2019 Apr 11;8(1):1.

9. Hu X, Daly M. What have we learned from six years of GWAS in autoimmune diseases, and what is next? Current Opinion in Immunology. Elsevier Current Trends; 2012 Oct 1;24(5):571–5.

10. Euesden J, Danese A, Lewis CM, Maughan B. A bidirectional relationship between depression and the autoimmune disorders - New perspectives from the National Child Development Study. PLOS ONE. Public Library of Science; 2017 Mar;12(3):1–14.

11. Wray NR, Ripke S, Mattheisen M, Trzaskowski M, Byrne EM, Abdellaoui A, et al. Genome-wide association analyses identify 44 risk variants and refine the genetic architecture of major depression. Nature Genetics. Nature Publishing Group; 2018 May;50:668–81.

12. Pouget JG, Schizophrenia Working Group of the Psychiatric Genomics Consortium, Han B, Wu Y, Mignot E, Ollila HM, et al. Cross-disorder analysis of schizophrenia and 19 immune-mediated diseases identifies shared genetic risk. Human Molecular Genetics. 2019 Jun 18.

13. Bycroft C, Freeman C, Petkova D, Band G, Elliott LT, Sharp K, et al. The UK Biobank resource with deep phenotyping and genomic data. Nature. Nature Publishing Group; 2018 Oct 1;562(7726):203–9.

14. World Health Organization. International Classification of Diseases (10th Edition). Geneva: World Health Organisation; 1992.

15. Davis KAS, Coleman JRI, Adams M, Allen N, Breen G, Cullen B, et al. Mental health in UK Biobank – development, implementation and results from an online questionnaire completed by 157 366 participants: a reanalysis. BJPsych Open. Cambridge University Press; 2020 Mar 1;6(2):e18.

16. Glanville KP, Coleman JRI, Howard DM, Pain O, Hanscombe KB, Jermy B, et al. Multiple measures of depression to enhance validity of major depressive disorder in the UK Biobank. BJPsych Open. Cambridge University Press; 2021 Mar 1;7(2):82.

17. Smith DJ, Nicholl BI, Cullen B, Martin D, Ul-Haq Z, Evans J, et al. Prevalence and Characteristics of Probable Major Depression and Bipolar Disorder within UK Biobank: Cross-Sectional Study of 172,751 Participants. Potash JB, editor. PLOS ONE. 2013 Nov 25;8(11):e75362.

18. Chang CC, Chow CC, Tellier LC, Vattikuti S, Purcell SM, Lee JJ. Second-generation PLINK: rising to the challenge of larger and richer datasets. GigaScience [Internet]. 2015 Feb 25;4(1):559. Available from: www.cog-genomics.org/plink/1.9/

19. Manichaikul A, Mychaleckyj JC, Rich SS, Daly K, Sale M, Chen W-M. Robust relationship inference in genome-wide association studies. Bioinformatics. 2010 Nov 15;26(22):2867–73.

20. Choi SW. choishingwan/GreedyRelated: Function update. 2020 Mar 4.

21. Abraham G, Qiu Y, Inouye M. FlashPCA2: principal component analysis of Biobank-scale genotype datasets. Bioinformatics. 2017 Sep 1;33(17):2776–8.

22. R Development Core Team. R: A language and environment for statistical computing [Internet]. Vienna, Austria: R Foundation for Statistical Computing; 2008. Available from: http://www.R-project.org

23. Dubois PCA, Trynka G, Franke L, Hunt KA, Romanos J, Curtotti A, et al. Multiple common variants for celiac disease influencing immune gene expression. Nature Genetics. Nature Publishing Group; 2010 Apr 1;42(4):295–302.

24. Liu JZ, van Sommeren S, Huang H, Ng SC, Alberts R, Takahashi A, et al. Association analyses identify 38 susceptibility loci for inflammatory bowel disease and highlight shared genetic risk across populations. Nature Genetics. Nature Publishing Group; 2015 Sep 1;47(9):979–86.

25. The International Multiple Sclerosis Genetics Consortium & The Wellcome Trust Case Control Consortium 2. Genetic risk and a primary role for cell-mediated immune mechanisms in multiple sclerosis. Nature. Nature Publishing Group; 2011 Aug 1;476(7359):214–9.

26. Renton AE, Pliner HA, Provenzano C, Evoli A, Ricciardi R, Nalls MA, et al. A Genome-Wide Association Study of Myasthenia Gravis. JAMA neurology. JAMA Neurol; 2015 Apr 1;72(4):396–404.

27. Tsoi LC, Stuart PE, Tian C, Gudjonsson JE, Das S, Zawistowski M, et al. Large scale meta-analysis characterizes genetic architecture for common psoriasis associated variants. Nature Communications. Nature Publishing Group; 2017 May 24;8(1):1–8.

28. Aterido A, Cañete JD, Tornero J, Ferrándiz C, Pinto JA, Gratacós J, et al. Genetic variation at the glycosaminoglycan metabolism pathway contributes to the risk of psoriatic arthritis but not psoriasis. Annals of the Rheumatic Diseases. BMJ Publishing Group Ltd; 2019 Mar 1;78(3):355–64.

29. Okada Y, Di Wu, Trynka G, Raj T, Terao C, Ikari K, et al. Genetics of rheumatoid arthritis contributes to biology and drug discovery. Nature. Nature Publishing Group; 2014 Feb 1;506(7488):376–81.

30. Bentham J, Morris DL, Graham DSC, Pinder CL, Tombleson P, Behrens TW, et al. Genetic association analyses implicate aberrant regulation of innate and adaptive immunity genes in the pathogenesis of systemic lupus erythematosus. Nature Genetics. Nature Publishing Group; 2015 Dec 1;47(12):1457–64.

31. Choi SW, O’Reilly PF. PRSice-2: Polygenic Risk Score software for biobank-scale data.- PubMed - NCBI. GigaScience. 2019 Jul 15;8(7):2091.

32. Andres E, Serraj K. Optimal management of pernicious anemia. Journal of blood medicine. J Blood Med; 2012;3:97–103.

33. Simmonds MJ, Gough SCL. Unravelling the genetic complexity of autoimmune thyroid disease: HLA, CTLA-4 and beyond. Clinical and Experimental Immunology. Wiley-Blackwell; 2004 Apr 1;136(1):1–10.

34. Bradfield JP, Qu H-Q, Wang K, Zhang H, Sleiman PM, Kim CE, et al. A Genome-Wide Meta-Analysis of Six Type 1 Diabetes Cohorts Identifies Multiple Associated Loci. McCarthy MI, editor. PLoS Genet. Public Library of Science; 2011 Sep 29;7(9):e1002293.

35. Spillane J, Higham E, Kullmann DM. Myasthenia gravis. BMJ. British Medical Journal Publishing Group; 2012 Dec 21;345(dec 21 3):e8497–7.

36. Tsoi LC, Stuart PE, Tian C, Gudjonsson JE, Das S, Zawistowski M, et al. Large scale meta-analysis characterizes genetic architecture for common psoriasis associated variants. - PubMed - NCBI. Nature Communications. 2017 May 24;8:15382.

37. International Genetics of Ankylosing Spondylitis Consortium (IGAS). Identification of multiple risk variants for ankylosing spondylitis through high-density genotyping of immune-related loci. Nature Genetics. Nature Publishing Group; 2013 Jul 1;45(7):730–8.

38. Partington RJ, Muller S, Helliwell T, Mallen CD, Sultan AA. Incidence, prevalence and treatment burden of polymyalgia rheumatica in the UK over two decades: a population- based study. Annals of the Rheumatic Diseases. BMJ Publishing Group Ltd; 2018 Dec 1;77(12):1750–6.

39. Humphreys JH, Verstappen SMM, Hyrich KL, Chipping JR, Marshall T, Symmons DPM. The incidence of rheumatoid arthritis in the UK: comparisons using the 2010 ACR/EULAR classification criteria and the 1987 ACR classification criteria. Results from the Norfolk Arthritis Register. Annals of the Rheumatic Diseases. BMJ Publishing Group Ltd; 2013 Aug 1;72(8):1315–20.

40. Lessard CJ, Li H, Adrianto I, Ice JA, Rasmussen A, Grundahl KM, et al. Variants at multiple loci implicated in both innate and adaptive immune responses are associated with Sjögren’s syndrome. Nature Genetics. Nat Genet; 2013 Oct 6;45(11):1284–92.

41. Rees F, Doherty M, Grainge M, Davenport G, Lanyon P, Zhang W. The incidence and prevalence of systemic lupus erythematosus in the UK, 1999–2012. Annals of the Rheumatic Diseases. BMJ Publishing Group Ltd; 2016 Jan 1;75(1):136–41.

42. Luigi Palla FD. A Fast Method that Uses Polygenic Scores to Estimate the Variance Explained by Genome-wide Marker Panels and the Proportion of Variants Affecting a Trait. Am J Hum Genet. Elsevier; 2015 Aug 6;97(2):250–9.

43. Bulik-Sullivan BK, Loh P-R, Finucane HK, Ripke S, Yang J, Schizophrenia Working Group of the Psychiatric Genomics Consortium, et al. LD Score regression distinguishes confounding from polygenicity in genome-wide association studies. Nature Genetics. 2015 Mar;47(3):291–5.

44. Gold SM, Köhler-Forsberg O, Moss-Morris R, Mehnert A, Miranda JJ, Bullinger M, et al. Comorbid depression in medical diseases. Nat Rev Dis Primers. Nature Publishing Group; 2020 Aug 20;6(1):1–22.

45. ME L, L T, J B, J C, de Boer P, Mondelli V, et al. Peripheral Blood Cell-Stratified Subgroups of Inflamed Depression. Biological Psychiatry. Biol Psychiatry; 2020 Jul 15;88(2):185–96.

46. Dube SR, Fairweather D, Pearson WS, Felitti VJ, Anda RF, Croft JB. Cumulative Childhood Stress and Autoimmune Diseases in Adults. Psychosomatic Medicine. 2009 Feb;71(2):243–50.

47. Hughes K, Bellis MA, Hardcastle KA, Sethi D, Butchart A, Mikton C, et al. The effect of multiple adverse childhood experiences on health: a systematic review and meta-analysis. The Lancet Public Health. Elsevier; 2017 Aug 1;2(8):e356–66.

48. A F, TJ L, C S, N D, L A, T S, et al. Comparison of Sociodemographic and Health-Related Characteristics of UK Biobank Participants With Those of the General Population. American Journal of Epidemiology. Am J Epidemiol; 2017 Nov 1;186(9):1026–34.

49. Batty GD, Gale C, Kivimaki M, Deary I, Bell S. Generalisability of Results from UK Biobank: Comparison With a Pooling of 18 Cohort Studies. medRxiv. Cold Spring Harbor Laboratory Press; 2019 Aug 13;:19004705.

50. Duncan L, Shen H, Gelaye B, Meijsen J, Ressler K, Feldman M, et al. Analysis of polygenic risk score usage and performance in diverse human populations. Nature Communications. Nature Publishing Group; 2019 Jul 25;10(1):1–9.

