## Supplementary for "Investigating pleiotropy between depression and autoimmune diseases using the UK Biobank"

#### *Supplement*

### 1 Power calculations

#### 1.1 SNP-based heritability estimates

Estimates of SNP-based heritability are required for power calculations using the AVENGEME software. We estimated SNP-based heritability for depression and the eight autoimmune diseases for which summary statistics were publicly available (summary statistics described in Table 1 of the main paper), and the results are shown in Supplementary Figure 1.

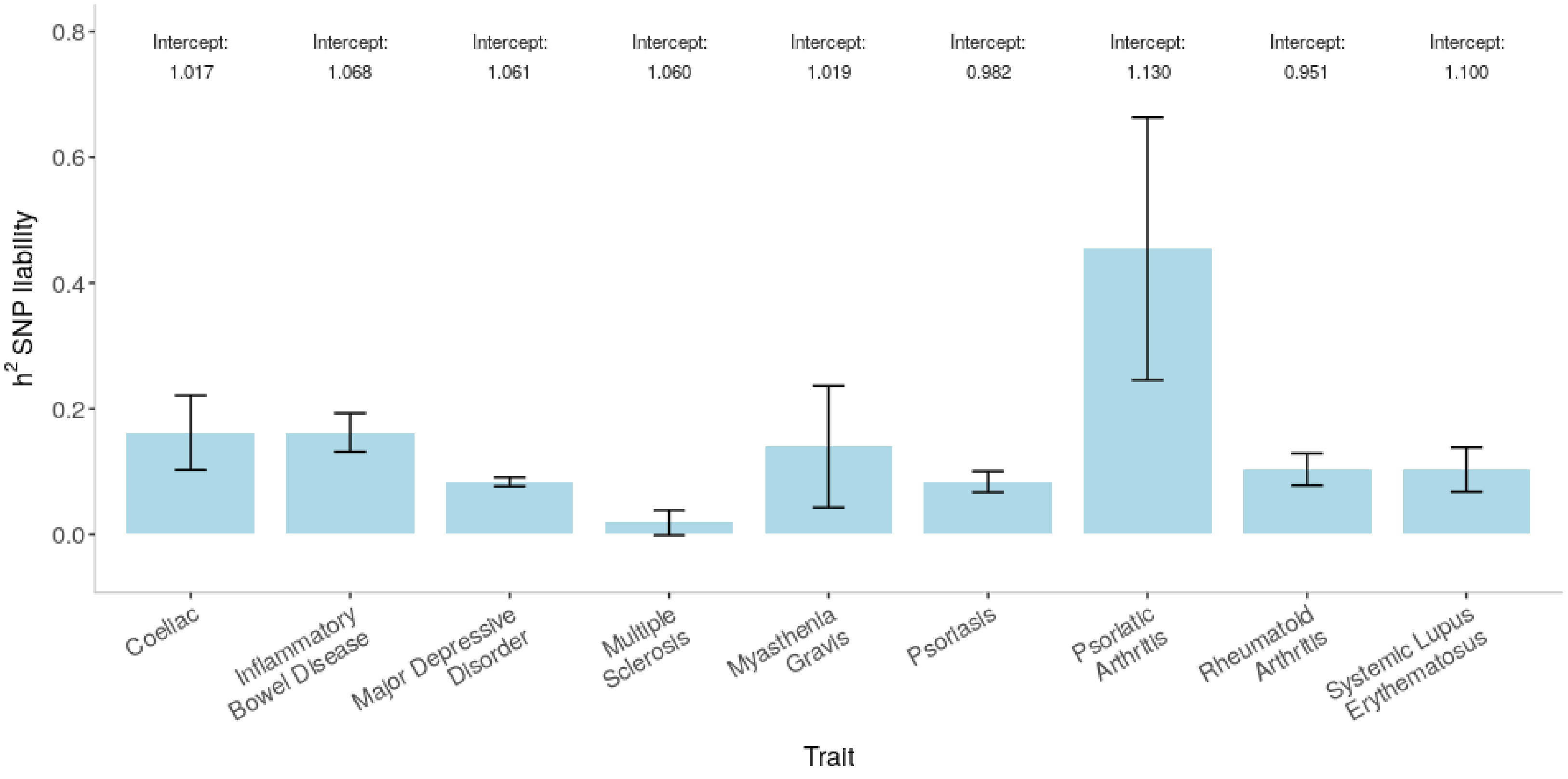

**Supplementary Figure 1**: SNP-based heritability ($h^2_{SNP}$) of traits with publicly available summary statistics. Estimated using LDSC after removing variants within the MHC from summary statistics. Transformed to liability scale using population prevalence estimates: Coeliac=1%, IBD=0.5%, MDD=15%, MS=0.1%, MG=0.02%, Psoriasis=2%, PsA=0.5%, RA=1%, SLE=0.1%. Error bars: 95% CIs. Intercepts shown atop bars.

#### 1.2 Power estimates for autoimmune PRS predicting any depression in the UKB

**Supplementary Table 1**: AVENGEME inputs for calculating power to detect association between PRS for autoimmune diseases and 'any' depression case status in the UKB.

| BASE GWAS | | | | | | | TARGET TRAIT | | | | |
|---|---|---|---|---|---|---|---|---|---|---|---|
| Trait | Total N (case+control) | $h^2_{SNP}$ excluding MHC | Population prevalence | Sample prevalence | Optimal p-value threshold ($P_T$) | No. SNPs used to construct PRS at ($P_T$) | Trait | Total N (case+control) | $h^2_{SNP}$ excluding MHC | Population prevalence | Sample prevalence |
| Coeliac | 15,283 | 16.21% | 1.00% | 30% | 0.200 | 12,430 | Any Depression | 297,627 | 8.38% | 15% | 22% |
| Inflammatory Bowel Disease | 34,652 | 16.20% | 0.50% | 37% | 1.000 | 80,201 | Any Depression | 297,627 | 8.38% | 15% | 22% |
| Multiple Sclerosis | 27,148 | 1.85% | 0.10% | 36% | 0.050 | 3,240 | Any Depression | 297,627 | 8.38% | 15% | 22% |
| Myasthenia Gravis | 3,030 | 13.96% | 0.02% | 34% | 0.300 | 56,894 | Any Depression | 297,627 | 8.38% | 15% | 22% |
| Psoriasis | 305,801 | 8.38% | 2.00% | 6% | 1.000 | 136,084 | Any Depression | 297,627 | 8.38% | 15% | 22% |
| Psoriatic Arthritis | 2,847 | 45.44% | 0.50% | 50% | 0.050 | 4,666 | Any Depression | 297,627 | 8.38% | 15% | 22% |
| Rheumatoid Arthritis | 58,284 | 10.35% | 1.00% | 25% | 1.000 | 118,649 | Any Depression | 297,627 | 8.38% | 15% | 22% |
| Systemic Lupus Erythematosus | 23,210 | 10.30% | 0.10% | 31% | 0.001 | 836 | Any Depression | 297,627 | 8.38% | 15% | 22% |

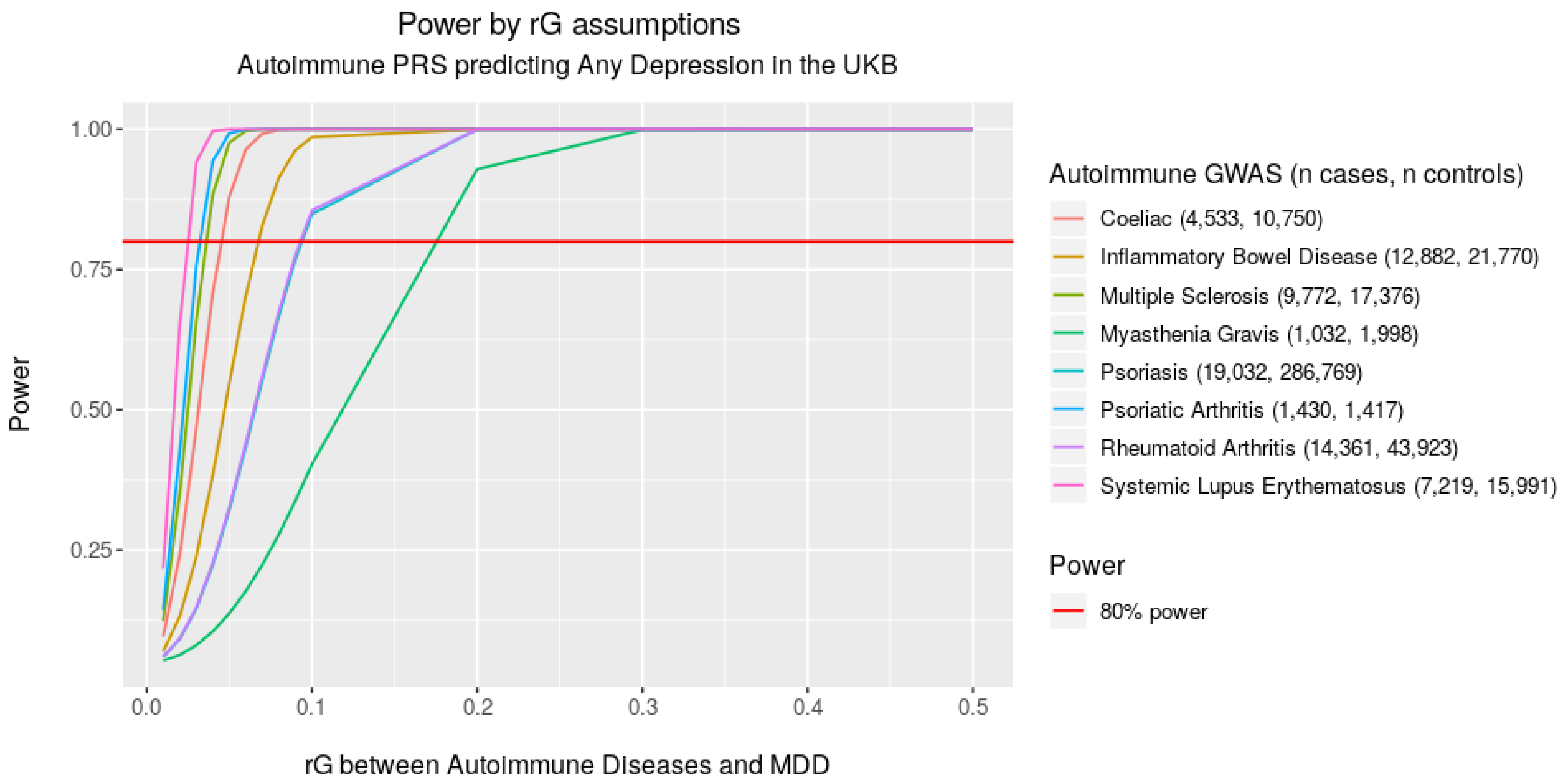

**Supplementary Figure 2**: Power to detect associations between autoimmune PRS and any depression case status in UKB across different levels of genetic correlation (rG).

#### 1.3 Power estimates for depression PRS predicting autoimmune disease in the UKB

**Supplementary Table 2**: AVENGEME inputs for calculating power to detect association between PRS for Major Depressive Disorder (MDD) and possible autoimmune diseases in UKB.

| BASE GWAS | | | | | | | TARGET TRAIT | | | | |
|---|---|---|---|---|---|---|---|---|---|---|---|
| Trait | Total N (case+control) | $h^2_{SNP}$ excluding MHC | Population prevalence | Sample prevalence | Optimal p-value threshold ($P_T$) | No. SNPs used to construct PRS at ($P_T$) | Trait | Total N (case+control) | $h^2_{SNP}$ excluding MHC | Population prevalence | Sample prevalence |
| MDD | 431,394 | 8.38% | 15% | 27% | 0.001 | 1,123 | Coeliac | 326,438 | 16.21% | 1.00% | 0.72% |
| MDD | 431,394 | 8.38% | 15% | 27% | 0.300 | 48,852 | Inflammatory Bowel Disease | 329,179 | 16.20% | 0.50% | 1.55% |
| MDD | 431,394 | 8.38% | 15% | 27% | 0.500 | 67,956 | Multiple Sclerosis | 325,757 | 1.85% | 0.10% | 0.52% |
| MDD | 431,394 | 8.38% | 15% | 27% | 1.000 | 99,216 | Myasthenia Gravis | 324,308 | 13.96% | 0.02% | 0.07% |
| MDD | 431,394 | 8.38% | 15% | 27% | 0.500 | 67,956 | Psoriasis | 329,533 | 8.38% | 2.00% | 1.66% |
| MDD | 431,394 | 8.38% | 15% | 27% | 1.000 | 99,216 | Psoriatic Arthritis | 325,181 | 45.44% | 0.50% | 0.34% |
| MDD | 431,394 | 8.38% | 15% | 27% | 0.500 | 67,956 | Rheumatoid Arthritis | 330,434 | 10.35% | 1.00% | 1.92% |
| MDD | 431,394 | 8.38% | 15% | 27% | 0.050 | 14,137 | Systemic Lupus Erythematosus | 324,698 | 10.30% | 0.10% | 0.19% |

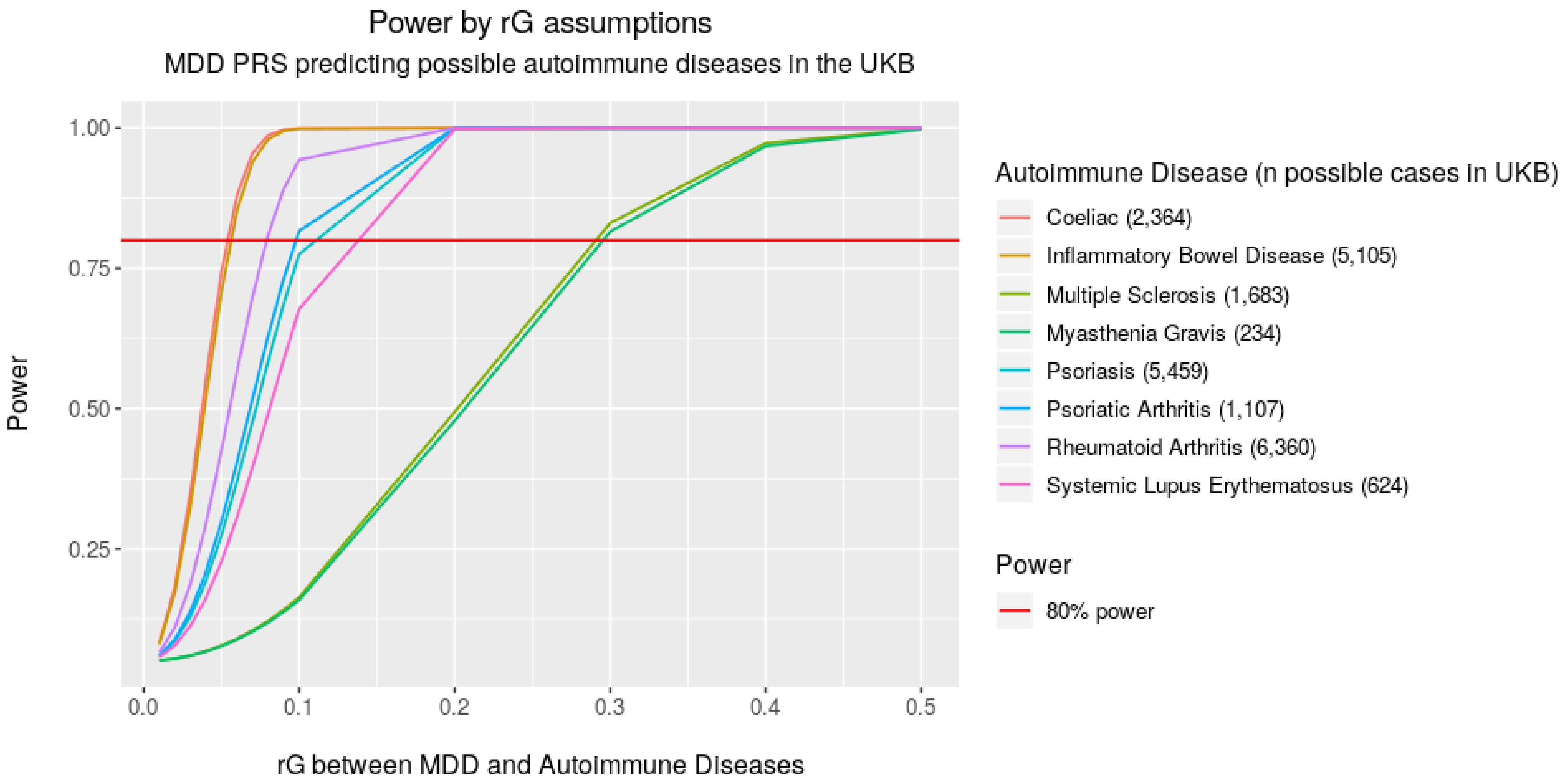

**Supplementary Figure 3:** Power to detect associations between MDD PRS and possible autoimmune case status in UKB across different levels of genetic correlation (rG).

### 2 Polygenic risk score analyses

**Supplementary Table 3**: Within-trait PRS - associations between PRS for autoimmune diseases and autoimmune case/control status (possible/probable) at optimal $P_T$.

| Base GWAS | Target Trait in UKB | Optimal p-value threshold ($P_T$) | No. SNPs used to construct PRS at ($P_T$) | Population Prevalence used to convert to liability $R^2$ | Unadjusted Variance explained (Observed $R^2$) | Adjusted Variance explained (Liability $R^2$) | Coefficient | Standard Error | P-value |
|---|---|---|---|---|---|---|---|---|---|
| Coeliac | Coeliac (Possible) | 1.000 | 42,731 | 1.00% | 1.65% | 2.60% | 0.46 | 0.022 | 2e-98 |
| Coeliac | Coeliac (Probable) | 1.000 | 42,731 | 1.00% | 1.91% | 3.37% | 0.52 | 0.030 | 6e-69 |
| Inflammatory Bowel Disease | Inflammatory Bowel Disease (Possible) | 0.001 | 676 | 0.50% | 2.41% | 2.77% | 0.49 | 0.014 | 5e-256 |
| Inflammatory Bowel Disease | Inflammatory Bowel Disease (Probable) | 0.001 | 676 | 0.50% | 2.90% | 3.62% | 0.56 | 0.017 | 2e-233 |
| Multiple Sclerosis | Multiple Sclerosis (Possible) | 0.001 | 230 | 0.10% | 1.42% | 1.53% | 0.42 | 0.025 | 4e-65 |
| Multiple Sclerosis | Multiple Sclerosis (Probable) | 0.001 | 230 | 0.10% | 1.43% | 1.64% | 0.43 | 0.030 | 2e-48 |
| Myasthenia Gravis | Myasthenia Gravis (Possible) | 0.100 | 22,895 | 0.02% | 0.31% | 0.35% | 0.23 | 0.066 | 6e-04 |
| Myasthenia Gravis | Myasthenia Gravis (Probable) | 0.100 | 22,895 | 0.02% | 0.28% | 0.34% | 0.23 | 0.084 | 7e-03 |
| Psoriasis | Psoriasis (Possible) | 0.001 | 637 | 2.00% | 2.08% | 3.24% | 0.44 | 0.013 | 5e-236 |
| Psoriasis | Psoriasis (Probable) | 0.001 | 637 | 2.00% | 2.27% | 4.11% | 0.49 | 0.019 | 1e-154 |
| Psoriatic Arthritis | Psoriatic Arthritis (Possible) | 0.200 | 14,198 | 0.50% | 0.19% | 0.29% | 0.27 | 0.052 | 2e-07 |
| Psoriatic Arthritis | Psoriatic Arthritis (Probable) | 1.000 | 45,914 | 0.50% | 0.20% | 0.34% | 0.31 | 0.066 | 3e-06 |
| Rheumatoid Arthritis | Rheumatoid Arthritis (Possible) | 0.001 | 550 | 1.00% | 0.58% | 0.73% | 0.23 | 0.013 | 3e-74 |
| Rheumatoid Arthritis | Rheumatoid Arthritis (Probable) | 0.001 | 550 | 1.00% | 1.12% | 1.63% | 0.34 | 0.017 | 2e-90 |
| Systemic Lupus Erythematosus | Systemic Lupus Erythematosus (Possible) | 0.001 | 836 | 0.10% | 1.37% | 1.73% | 0.42 | 0.037 | 6e-30 |
| Systemic Lupus Erythematosus | Systemic Lupus Erythematosus (Probable) | 0.300 | 74,372 | 0.10% | 2.14% | 2.92% | 0.58 | 0.052 | 6e-29 |

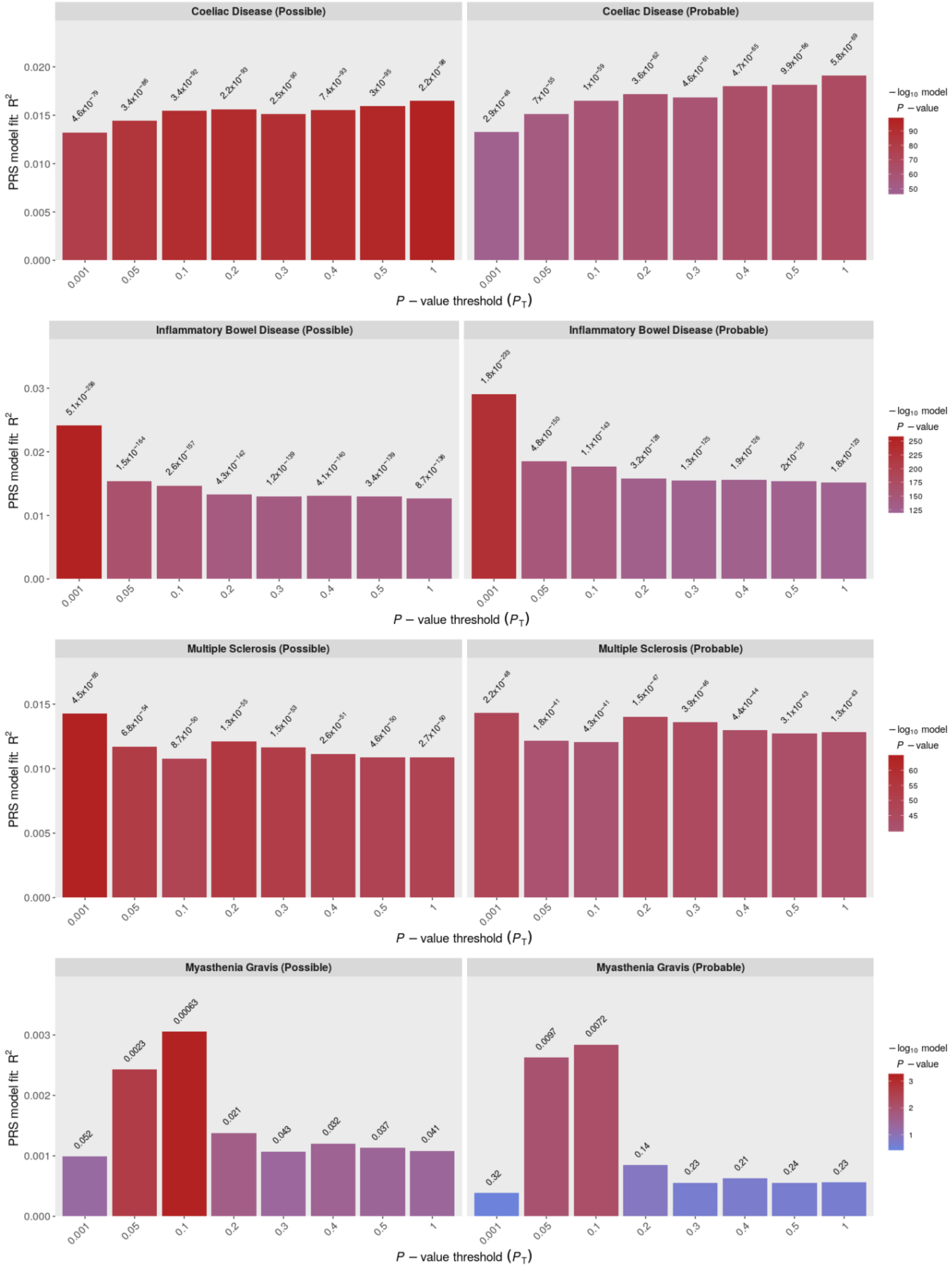

Coeliac Disease (Possible)
Coeliac Disease (Probable)
Inflammatory Bowel Disease (Possible)
Inflammatory Bowel Disease (Probable)
Multiple Sclerosis (Possible)
Multiple Sclerosis (Probable)
Myasthenia Gravis (Possible)
Myasthenia Gravis (Probable)
PRS model fit: R²
P − value threshold (P_T)
− log10 model P − value
0.001
0.05
0.1
0.2
0.3
0.4
0.5
1
0.052
0.0023
0.00063
0.021
0.043
0.032
0.037
0.041
0.32
0.0097
0.0072
0.14
0.23
0.21
0.24
0.23

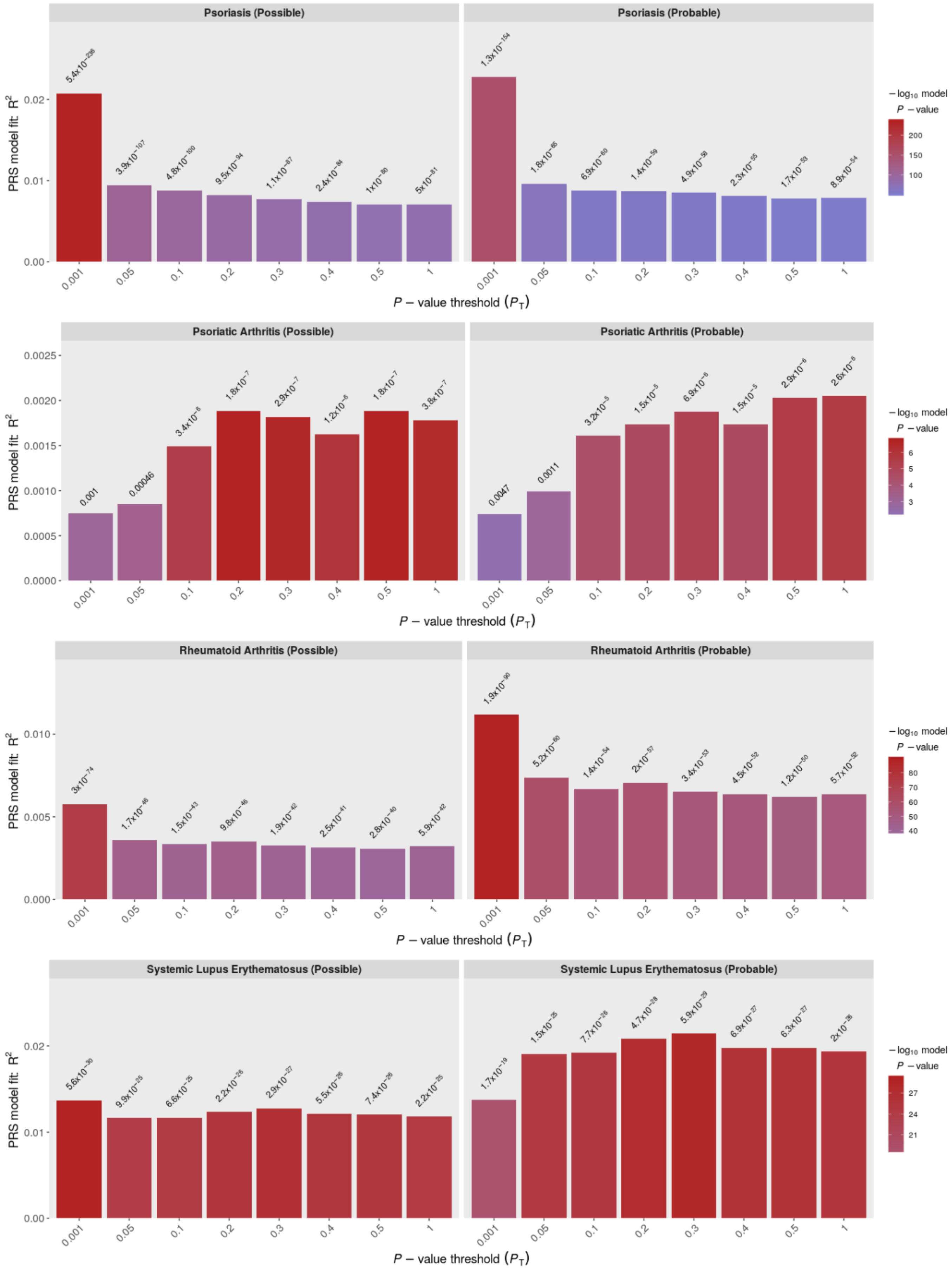

**Supplementary Figure 4:** Within-trait PRS - associations between PRS for autoimmune diseases and autoimmune case/control status (possible/probable) across eight $P_T$. Y-axis shows unadjusted $R^2$ (observed-scale).

**Supplementary Table 4**: Within-trait PRS - associations between PRS for MDD and depression case/control status (any/stringent) at optimal $P_T$.

| Base GWAS | Target Trait in UKB | Optimal p-value threshold ($P_T$) | No. SNPs used to construct PRS at ($P_T$) | Population Prevalence used to convert to liability $R^2$ | Unadjusted Variance explained (Observed $R^2$) | Adjusted Variance explained (Liability $R^2$) | Coefficient | Standard Error | P-value |
|---|---|---|---|---|---|---|---|---|---|
| Major Depressive Disorder | Depression (Any) | 0.4 | 59,001 | 15% | 1.30% | 1.48% | 0.23 | 0.005 | <5e-324 |
| Major Depressive Disorder | Depression (Stringent) | 0.4 | 59,001 | 15% | 1.16% | 2.23% | 0.28 | 0.009 | 2e-228 |

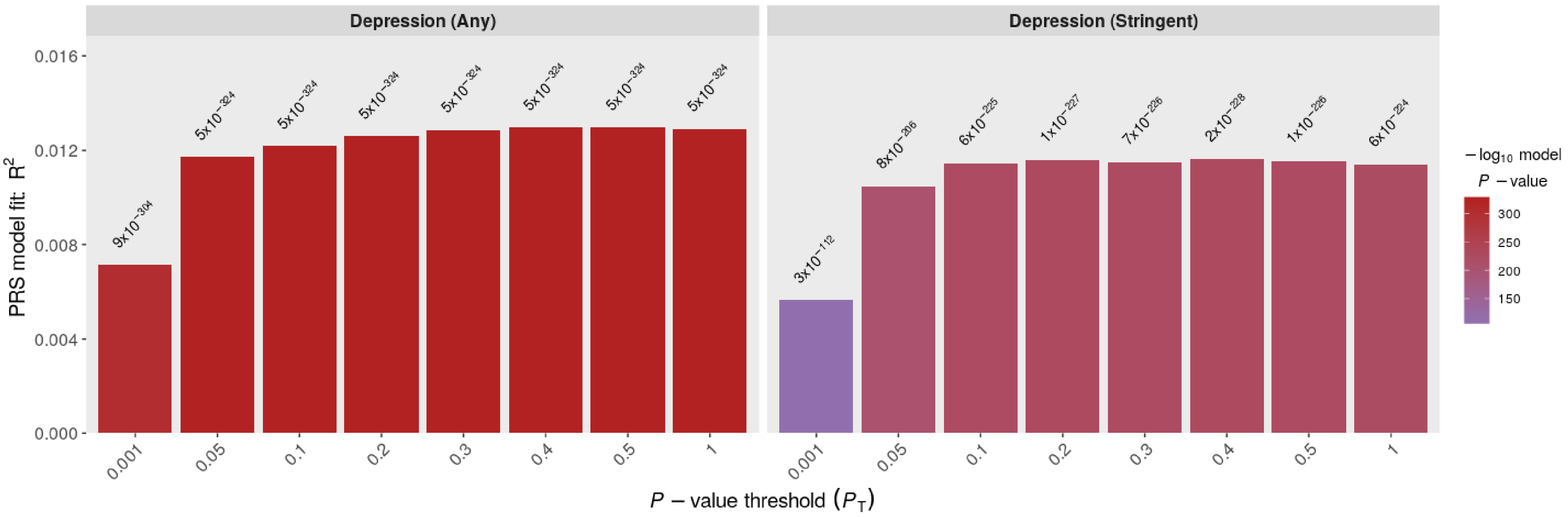

**Supplementary Figure 5**: Within-trait PRS - associations between PRS for MDD and depression case/control status (any/stringent) across eight $P_T$. Y-axis shows unadjusted $R^2$ (observed-scale). Note: p-value for MDD PRS predicting Depression (Any) = 0 for thresholds 0.05 to 1 due to limit on the smallest number on the R platform (5e-324).

**Supplementary Table 5**: Cross-trait PRS - associations between PRS for autoimmune diseases and depression case/control status (any/stringent) in men and women at optimal $P_T$. Associations with p-values < 7.1x10-5, meeting Bonferroni correction, highlighted in red.

| Base GWAS | Target Trait in UKB | Optimal p-value threshold ($P_T$) | No. SNPs used to construct PRS at ($P_T$) | Population Prevalence used to convert to liability $R^2$ | Unadjusted Variance explained (Observed $R^2$) | Adjusted Variance explained (Liability $R^2$) | Coefficient | Standard Error | P-value |
|---|---|---|---|---|---|---|---|---|---|
| Coeliac | Any (combined) | 0.200 | 12,430 | 15% | 0.0021% | 0.0024% | 0.009 | 0.005 | 4.4e-02 |
| Coeliac | Any (female) | 0.200 | 12,430 | 15% | 0.0006% | 0.0007% | 0.005 | 0.006 | 4.1e-01 |
| Coeliac | Any (male) | 0.050 | 3,881 | 15% | 0.0045% | 0.0060% | 0.015 | 0.008 | 5.4e-02 |
| Coeliac | Stringent (combined) | 0.200 | 12,430 | 15% | 0.0013% | 0.0026% | 0.010 | 0.009 | 2.7e-01 |
| Coeliac | Stringent (female) | 0.001 | 186 | 15% | 0.0017% | 0.0029% | 0.010 | 0.011 | 3.4e-01 |
| Coeliac | Stringent (male) | 0.001 | 186 | 15% | 0.0034% | 0.0075% | -0.016 | 0.015 | 2.7e-01 |
| Inflammatory Bowel Disease | Any (combined) | 1.000 | 80,201 | 15% | 0.0051% | 0.0058% | 0.014 | 0.005 | 1.6e-03 |
| Inflammatory Bowel Disease | Any (female) | 1.000 | 80,201 | 15% | 0.0047% | 0.0049% | 0.013 | 0.006 | 2.4e-02 |
| Inflammatory Bowel Disease | Any (male) | 1.000 | 80,201 | 15% | 0.0079% | 0.0106% | 0.019 | 0.008 | 1.0e-02 |
| Inflammatory Bowel Disease | Stringent (combined) | 0.300 | 38,677 | 15% | 0.0173% | 0.0333% | 0.035 | 0.009 | 7.9e-05 |
| Inflammatory Bowel Disease | Stringent (female) | 0.050 | 10,082 | 15% | 0.0238% | 0.0406% | 0.038 | 0.011 | 4.4e-04 |
| Inflammatory Bowel Disease | Stringent (male) | 1.000 | 80,201 | 15% | 0.0230% | 0.0510% | 0.043 | 0.015 | 4.2e-03 |
| Lupus | Any (combined) | 0.001 | 836 | 15% | 0.0029% | 0.0034% | -0.011 | 0.004 | 1.7e-02 |
| Lupus | Any (female) | 0.001 | 836 | 15% | 0.0026% | 0.0027% | -0.010 | 0.006 | 9.4e-02 |
| Lupus | Any (male) | 0.001 | 836 | 15% | 0.0056% | 0.0075% | -0.016 | 0.007 | 3.1e-02 |
| Lupus | Stringent (combined) | 0.001 | 836 | 15% | 0.0018% | 0.0035% | -0.011 | 0.009 | 2.0e-01 |
| Lupus | Stringent (female) | 0.001 | 836 | 15% | 0.0038% | 0.0066% | -0.015 | 0.011 | 1.6e-01 |
| Lupus | Stringent (male) | 0.050 | 18,572 | 15% | 0.0020% | 0.0045% | 0.013 | 0.015 | 3.9e-01 |
| Multiple Sclerosis | Any (combined) | 0.050 | 3,240 | 15% | 0.0052% | 0.0059% | 0.014 | 0.005 | 1.5e-03 |
| Multiple Sclerosis | Any (female) | 0.050 | 3,240 | 15% | 0.0066% | 0.0068% | 0.015 | 0.006 | 7.4e-03 |
| Multiple Sclerosis | Any (male) | 0.050 | 3,240 | 15% | 0.0058% | 0.0078% | 0.016 | 0.007 | 2.8e-02 |
| Multiple Sclerosis | Stringent (combined) | 0.050 | 3,240 | 15% | 0.0033% | 0.0063% | 0.015 | 0.009 | 8.6e-02 |
| Multiple Sclerosis | Stringent (female) | 0.100 | 5,974 | 15% | 0.0087% | 0.0148% | 0.023 | 0.011 | 3.4e-02 |
| Multiple Sclerosis | Stringent (male) | 0.050 | 3,240 | 15% | 0.0024% | 0.0053% | 0.014 | 0.015 | 3.6e-01 |
| Myasthenia Gravis | Any (combined) | 0.300 | 56,894 | 15% | 0.0084% | 0.0096% | 0.018 | 0.005 | 5.2e-05 |
| Myasthenia Gravis | Any (female) | 0.200 | 40,875 | 15% | 0.0127% | 0.0131% | 0.022 | 0.006 | 2.1e-04 |
| Myasthenia Gravis | Any (male) | 0.300 | 56,894 | 15% | 0.0073% | 0.0098% | 0.019 | 0.008 | 1.4e-02 |
| Myasthenia Gravis | Stringent (combined) | 0.300 | 56,894 | 15% | 0.0208% | 0.0399% | 0.038 | 0.009 | 1.6e-05 |
| Myasthenia Gravis | Stringent (female) | 0.300 | 56,894 | 15% | 0.0164% | 0.0279% | 0.032 | 0.011 | 3.5e-03 |
| Myasthenia Gravis | Stringent (male) | 0.300 | 56,894 | 15% | 0.0384% | 0.0849% | 0.055 | 0.015 | 2.2e-04 |

| Base GWAS | Target Trait in UKB | Optimal p-value threshold ($P_T$) | No. SNPs used to construct PRS at ($P_T$) | Population Prevalence used to convert to liability $R^2$ | Unadjusted Variance explained (Observed $R^2$) | Adjusted Variance explained (Liability $R^2$) | Coefficient | Standard Error | P-value |
|---|---|---|---|---|---|---|---|---|---|
| Psoriasis | Any (combined) | 1.000 | 136,084 | 15% | 0.0101% | 0.0116% | 0.020 | 0.005 | 8.7e-06 |
| Psoriasis | Any (female) | 1.000 | 136,084 | 15% | 0.0124% | 0.0128% | 0.021 | 0.006 | 2.5e-04 |
| Psoriasis | Any (male) | 0.050 | 13,899 | 15% | 0.0133% | 0.0178% | 0.025 | 0.007 | 8.9e-04 |
| Psoriasis | Stringent (combined) | 0.050 | 13,899 | 15% | 0.0126% | 0.0243% | 0.029 | 0.009 | 7.6e-04 |
| Psoriasis | Stringent (female) | 1.000 | 136,084 | 15% | 0.0177% | 0.0303% | 0.033 | 0.011 | 2.4e-03 |
| Psoriasis | Stringent (male) | 0.050 | 13,899 | 15% | 0.0093% | 0.0207% | 0.027 | 0.015 | 6.8e-02 |
| Psoriatic Arthritis | Any (combined) | 0.050 | 4,666 | 15% | 0.0013% | 0.0015% | -0.010 | 0.007 | 1.1e-01 |
| Psoriatic Arthritis | Any (female) | 1.000 | 45,914 | 15% | 0.0026% | 0.0027% | 0.018 | 0.011 | 9.2e-02 |
| Psoriatic Arthritis | Any (male) | 0.100 | 8,135 | 15% | 0.0058% | 0.0077% | -0.026 | 0.012 | 2.8e-02 |
| Psoriatic Arthritis | Stringent (combined) | 0.001 | 211 | 15% | 0.0018% | 0.0036% | 0.012 | 0.009 | 2.0e-01 |
| Psoriatic Arthritis | Stringent (female) | 1.000 | 45,914 | 15% | 0.0086% | 0.0147% | 0.042 | 0.020 | 3.4e-02 |
| Psoriatic Arthritis | Stringent (male) | 0.100 | 8,135 | 15% | 0.0081% | 0.0178% | -0.039 | 0.023 | 9.1e-02 |
| Rheumatoid Arthritis | Any (combined) | 1.000 | 118,649 | 15% | 0.0006% | 0.0007% | 0.005 | 0.005 | 2.6e-01 |
| Rheumatoid Arthritis | Any (female) | 0.001 | 550 | 15% | 0.0011% | 0.0011% | -0.006 | 0.006 | 2.8e-01 |
| Rheumatoid Arthritis | Any (male) | 1.000 | 118,649 | 15% | 0.0059% | 0.0079% | 0.017 | 0.008 | 2.7e-02 |
| Rheumatoid Arthritis | Stringent (combined) | 1.000 | 118,649 | 15% | 0.0095% | 0.0183% | 0.025 | 0.009 | 3.4e-03 |
| Rheumatoid Arthritis | Stringent (female) | 0.001 | 550 | 15% | 0.0047% | 0.0080% | 0.017 | 0.011 | 1.2e-01 |
| Rheumatoid Arthritis | Stringent (male) | 1.000 | 118,649 | 15% | 0.0232% | 0.0514% | 0.043 | 0.015 | 4.1e-03 |

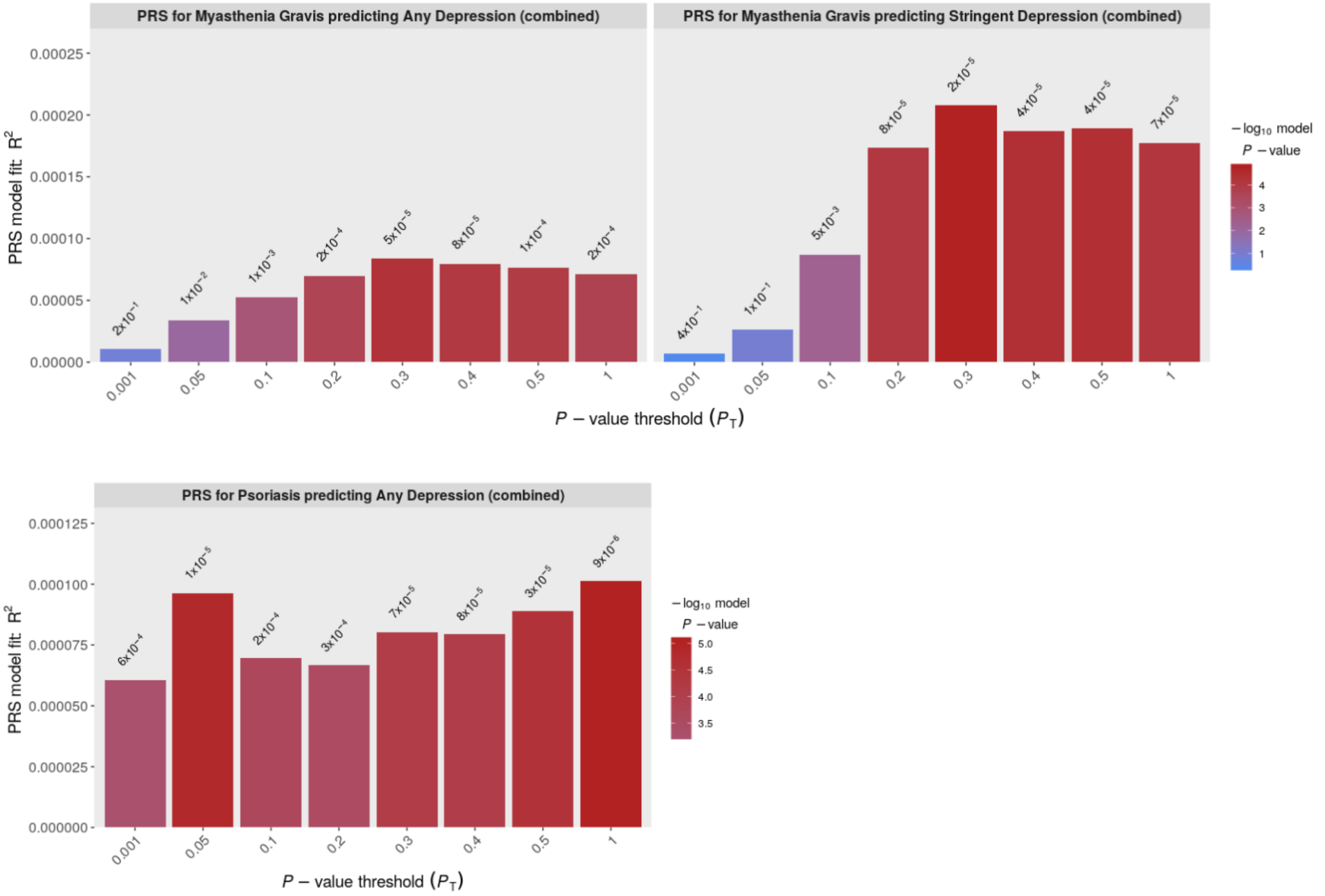

**Supplementary Figure 6**: Cross-trait PRS –associations between PRS for autoimmune diseases and depression case/control status (any/stringent) in men and women across eight $P_T$. Y-axis shows unadjusted $R^2$ (observed-scale). Bar charts are shown for significant results only.

**Supplementary Table 6**: Cross-trait PRS - associations between PRS for MDD and autoimmune case/control status (possible/probable) in men and women at optimal $P_T$. MDD = Major Depressive Disorder. Associations with p-values < 7.1x10$^{-5}$, meeting Bonferroni correction, highlighted in red. Sex*PRS p-value – estimated where significant associations were observed in sex-specific analyses; tested for an interaction between sex and PRS in the full sample (Phenotype ~ sex + PRS + sex*PRS + covariates).

| Base GWAS | Target Trait in UKB | Optimal p-value threshold ($P_T$) | No. SNPs used to construct PRS at ($P_T$) | Population Prevalence used to convert to liability $R^2$ | Unadjusted Variance explained (Observed $R^2$) | Adjusted Variance explained (Liability $R^2$) | Coefficient | Standard Error | P-value | Sex*PRS p-value |
|---|---|---|---|---|---|---|---|---|---|---|
| MDD | Ankylosing Spondylitis, Possible (combined) | 0.050 | 14,137 | 0.55% | 0.035% | 0.054% | 0.067 | 0.027 | 1.4e-02 | NA |
| MDD | Ankylosing Spondylitis, Possible (female) | 0.001 | 1,123 | 0.55% | 0.130% | 0.210% | 0.132 | 0.044 | 2.8e-03 | NA |
| MDD | Ankylosing Spondylitis, Possible (male) | 1.000 | 99,216 | 0.55% | 0.043% | 0.063% | 0.072 | 0.035 | 3.9e-02 | NA |
| MDD | Ankylosing Spondylitis, Probable (combined) | 0.001 | 1,123 | 0.55% | 0.003% | 0.006% | -0.022 | 0.049 | 6.5e-01 | NA |
| MDD | Ankylosing Spondylitis, Probable (female) | 0.400 | 59,001 | 0.55% | 0.032% | 0.064% | -0.074 | 0.096 | 4.5e-01 | NA |
| MDD | Ankylosing Spondylitis, Probable (male) | 0.001 | 1,123 | 0.55% | 0.005% | 0.008% | -0.026 | 0.057 | 6.5e-01 | NA |
| MDD | Autoimmune Thyroid Disease, Possible (combined) | 0.500 | 67,956 | 2.00% | 0.078% | 0.173% | 0.104 | 0.034 | 2.6e-03 | NA |
| MDD | Autoimmune Thyroid Disease, Possible (female) | 0.500 | 67,956 | 2.00% | 0.075% | 0.154% | 0.098 | 0.037 | 8.8e-03 | NA |
| MDD | Autoimmune Thyroid Disease, Possible (male) | 0.500 | 67,956 | 2.00% | 0.110% | 0.278% | 0.133 | 0.087 | 1.3e-01 | NA |
| MDD | Autoimmune Thyroid Disease, Probable (combined) | 0.400 | 59,001 | 2.00% | 0.083% | 0.193% | 0.110 | 0.041 | 7.3e-03 | NA |
| MDD | Autoimmune Thyroid Disease, Probable (female) | 0.400 | 59,001 | 2.00% | 0.082% | 0.178% | 0.105 | 0.044 | 1.7e-02 | NA |
| MDD | Autoimmune Thyroid Disease, Probable (male) | 0.001 | 1,123 | 2.00% | 0.138% | 0.363% | -0.153 | 0.108 | 1.5e-01 | NA |
| MDD | Coeliac, Possible (combined) | 0.001 | 1,123 | 1.00% | 0.109% | 0.172% | -0.112 | 0.021 | 6.0e-08 | NA |
| MDD | Coeliac, Possible (female) | 0.001 | 1,123 | 1.00% | 0.147% | 0.221% | -0.127 | 0.025 | 5.4e-07 | 0.33 |
| MDD | Coeliac, Possible (male) | 0.001 | 1,123 | 1.00% | 0.054% | 0.091% | -0.082 | 0.036 | 2.3e-02 | NA |
| MDD | Coeliac, Probable (combined) | 0.001 | 1,123 | 1.00% | 0.059% | 0.104% | -0.087 | 0.028 | 2.0e-03 | NA |
| MDD | Coeliac, Probable (female) | 0.001 | 1,123 | 1.00% | 0.066% | 0.110% | -0.090 | 0.034 | 8.7e-03 | NA |
| MDD | Coeliac, Probable (male) | 0.001 | 1,123 | 1.00% | 0.048% | 0.090% | -0.081 | 0.050 | 1.1e-01 | NA |

| Base GWAS | Target Trait in UKB | Optimal p-value threshold ($P_T$) | No. SNPs used to construct PRS at ($P_T$) | Population Prevalence used to convert to liability $R^2$ | Unadjusted Variance explained (Observed $R^2$) | Adjusted Variance explained (Liability $R^2$) | Coefficient | Standard Error | P-value | Sex*PRS p-value |
|---|---|---|---|---|---|---|---|---|---|---|
| MDD | Inflammatory Bowel Disease, Possible (combined) | 0.300 | 48,852 | 0.50% | 0.051% | 0.058% | 0.071 | 0.014 | 6.5e-07 | NA |
| MDD | Inflammatory Bowel Disease, Possible (female) | 0.050 | 14,137 | 0.50% | 0.103% | 0.119% | 0.100 | 0.020 | 4.0e-07 | 0.02 |
| MDD | Inflammatory Bowel Disease, Possible (male) | 0.300 | 48,852 | 0.50% | 0.044% | 0.051% | 0.066 | 0.020 | 1.2e-03 | NA |
| MDD | Inflammatory Bowel Disease, Probable (combined) | 0.050 | 14,137 | 0.50% | 0.029% | 0.036% | 0.055 | 0.017 | 1.2e-03 | NA |
| MDD | Inflammatory Bowel Disease, Probable (female) | 0.050 | 14,137 | 0.50% | 0.083% | 0.104% | 0.094 | 0.024 | 9.0e-05 | NA |
| MDD | Inflammatory Bowel Disease, Probable (male) | 1.000 | 99,216 | 0.50% | 0.014% | 0.017% | 0.038 | 0.024 | 1.1e-01 | NA |
| MDD | Multiple Sclerosis, Possible (combined) | 0.500 | 67,956 | 0.10% | 0.047% | 0.051% | 0.076 | 0.025 | 1.9e-03 | NA |
| MDD | Multiple Sclerosis, Possible (female) | 0.001 | 1,123 | 0.10% | 0.058% | 0.059% | 0.082 | 0.029 | 4.4e-03 | NA |
| MDD | Multiple Sclerosis, Possible (male) | 0.050 | 14,137 | 0.10% | 0.115% | 0.136% | 0.125 | 0.047 | 8.1e-03 | NA |
| MDD | Multiple Sclerosis, Probable (combined) | 1.000 | 99,216 | 0.10% | 0.064% | 0.073% | 0.091 | 0.030 | 2.0e-03 | NA |
| MDD | Multiple Sclerosis, Probable (female) | 0.001 | 1,123 | 0.10% | 0.081% | 0.088% | 0.100 | 0.035 | 3.9e-03 | NA |
| MDD | Multiple Sclerosis, Probable (male) | 0.050 | 14,137 | 0.10% | 0.160% | 0.201% | 0.152 | 0.057 | 7.6e-03 | NA |
| MDD | Myasthenia Gravis, Possible (combined) | 1.000 | 99,216 | 0.02% | 0.045% | 0.052% | 0.087 | 0.066 | 1.9e-01 | NA |
| MDD | Myasthenia Gravis, Possible (female) | 1.000 | 99,216 | 0.02% | 0.178% | 0.202% | 0.171 | 0.088 | 5.1e-02 | NA |
| MDD | Myasthenia Gravis, Possible (male) | 0.100 | 22,831 | 0.02% | 0.014% | 0.016% | -0.048 | 0.099 | 6.3e-01 | NA |
| MDD | Myasthenia Gravis, Probable (combined) | 0.001 | 1,123 | 0.02% | 0.014% | 0.017% | 0.050 | 0.082 | 5.5e-01 | NA |
| MDD | Myasthenia Gravis, Probable (female) | 1.000 | 99,216 | 0.02% | 0.075% | 0.092% | 0.115 | 0.119 | 3.3e-01 | NA |
| MDD | Myasthenia Gravis, Probable (male) | 1.000 | 99,216 | 0.02% | 0.071% | 0.085% | -0.112 | 0.116 | 3.4e-01 | NA |
| MDD | Pernicious Anemia, Possible (combined) | 0.050 | 14,137 | 0.10% | 0.035% | 0.038% | 0.066 | 0.025 | 9.4e-03 | NA |

| Base GWAS | Target Trait in UKB | Optimal p-value threshold ($P_T$) | No. SNPs used to construct PRS at ($P_T$) | Population Prevalence used to convert to liability $R^2$ | Unadjusted Variance explained (Observed $R^2$) | Adjusted Variance explained (Liability $R^2$) | Coefficient | Standard Error | P-value | Sex*PRS p-value |
|---|---|---|---|---|---|---|---|---|---|---|
| MDD | Pernicious Anemia, Possible (female) | 0.200 | 36,975 | 0.10% | 0.030% | 0.031% | 0.059 | 0.030 | 5.0e-02 | NA |
| MDD | Pernicious Anemia, Possible (male) | 0.500 | 67,956 | 0.10% | 0.059% | 0.070% | 0.090 | 0.048 | 6.0e-02 | NA |
| MDD | Pernicious Anemia, Probable (combined) | 0.050 | 14,137 | 0.10% | 0.113% | 0.150% | 0.131 | 0.049 | 7.2e-03 | NA |
| MDD | Pernicious Anemia, Probable (female) | 0.100 | 22,831 | 0.10% | 0.064% | 0.082% | 0.097 | 0.057 | 9.3e-02 | NA |
| MDD | Pernicious Anemia, Probable (male) | 0.050 | 14,137 | 0.10% | 0.380% | 0.544% | 0.250 | 0.093 | 6.9e-03 | NA |
| MDD | Polymyalgia Rheumatica/GCA, Possible (combined) | 0.200 | 36,975 | 0.85% | 0.058% | 0.095% | 0.085 | 0.025 | 6.8e-04 | NA |
| MDD | Polymyalgia Rheumatica/GCA, Possible (female) | 0.200 | 36,975 | 0.85% | 0.114% | 0.177% | 0.116 | 0.031 | 1.5e-04 | NA |
| MDD | Polymyalgia Rheumatica/GCA, Possible (male) | 0.001 | 1,123 | 0.85% | 0.007% | 0.012% | 0.030 | 0.043 | 4.8e-01 | NA |
| MDD | Polymyalgia Rheumatica/GCA, Probable (combined) | 0.200 | 36,975 | 0.85% | 0.074% | 0.132% | 0.100 | 0.034 | 2.8e-03 | NA |
| MDD | Polymyalgia Rheumatica/GCA, Probable (female) | 0.200 | 36,975 | 0.85% | 0.110% | 0.189% | 0.120 | 0.041 | 3.3e-03 | NA |
| MDD | Polymyalgia Rheumatica/GCA, Probable (male) | 0.001 | 1,123 | 0.85% | 0.113% | 0.214% | 0.128 | 0.059 | 3.0e-02 | NA |
| MDD | Psoriasis, Possible (combined) | 0.500 | 67,956 | 2.00% | 0.061% | 0.095% | 0.076 | 0.014 | 2.5e-08 | NA |
| MDD | Psoriasis, Possible (female) | 0.500 | 67,956 | 2.00% | 0.062% | 0.100% | 0.079 | 0.020 | 1.0e-04 | NA |
| MDD | Psoriasis, Possible (male) | 0.500 | 67,956 | 2.00% | 0.060% | 0.091% | 0.075 | 0.019 | 5.8e-05 | 0.86 |
| MDD | Psoriasis, Probable (combined) | 0.500 | 67,956 | 2.00% | 0.077% | 0.139% | 0.093 | 0.019 | 1.4e-06 | NA |
| MDD | Psoriasis, Probable (female) | 0.500 | 67,956 | 2.00% | 0.118% | 0.223% | 0.117 | 0.030 | 7.9e-05 | NA |
| MDD | Psoriasis, Probable (male) | 0.050 | 14,137 | 2.00% | 0.061% | 0.106% | 0.081 | 0.025 | 1.3e-03 | NA |
| MDD | Psoriatic Arthritis, Possible (combined) | 1.000 | 99,216 | 0.50% | 0.105% | 0.163% | 0.118 | 0.030 | 9.7e-05 | NA |
| MDD | Psoriatic Arthritis, Possible (female) | 1.000 | 99,216 | 0.50% | 0.159% | 0.248% | 0.145 | 0.043 | 6.5e-04 | NA |
| MDD | Psoriatic Arthritis, Possible (male) | 0.001 | 1,123 | 0.50% | 0.069% | 0.107% | 0.095 | 0.043 | 2.6e-02 | NA |
| MDD | Psoriatic Arthritis, Probable (combined) | 0.300 | 48,852 | 0.50% | 0.167% | 0.274% | 0.153 | 0.036 | 2.2e-05 | NA |

| Base GWAS | Target Trait in UKB | Optimal p-value threshold ($P_T$) | No. SNPs used to construct PRS at ($P_T$) | Population Prevalence used to convert to liability $R^2$ | Unadjusted Variance explained (Observed $R^2$) | Adjusted Variance explained (Liability $R^2$) | Coefficient | Standard Error | P-value | Sex*PRS p-value |
|---|---|---|---|---|---|---|---|---|---|---|
| MDD | Psoriatic Arthritis, Probable (female) | 1.000 | 99,216 | 0.50% | 0.212% | 0.349% | 0.173 | 0.051 | 6.4e-04 | NA |
| MDD | Psoriatic Arthritis, Probable (male) | 0.001 | 1,123 | 0.50% | 0.153% | 0.250% | 0.146 | 0.051 | 4.4e-03 | NA |
| MDD | Rheumatoid Arthritis, Possible (combined) | 0.500 | 67,956 | 1.00% | 0.065% | 0.082% | 0.078 | 0.013 | 1.1e-09 | NA |
| MDD | Rheumatoid Arthritis, Possible (female) | 0.300 | 48,852 | 1.00% | 0.060% | 0.071% | 0.072 | 0.016 | 3.5e-06 | 0.68 |
| MDD | Rheumatoid Arthritis, Possible (male) | 1.000 | 99,216 | 1.00% | 0.090% | 0.125% | 0.096 | 0.022 | 1.6e-05 | 0.26 |
| MDD | Rheumatoid Arthritis, Probable (combined) | 0.100 | 22,831 | 1.00% | 0.065% | 0.095% | 0.083 | 0.017 | 1.2e-06 | NA |
| MDD | Rheumatoid Arthritis, Probable (female) | 0.300 | 48,852 | 1.00% | 0.070% | 0.096% | 0.084 | 0.021 | 4.5e-05 | 0.90 |
| MDD | Rheumatoid Arthritis, Probable (male) | 1.000 | 99,216 | 1.00% | 0.075% | 0.121% | 0.094 | 0.032 | 2.9e-03 | NA |
| MDD | Sjögren Syndrome, Possible (combined) | 0.200 | 36,975 | 0.10% | 0.064% | 0.080% | 0.096 | 0.040 | 1.5e-02 | NA |
| MDD | Sjögren Syndrome, Possible (female) | 0.200 | 36,975 | 0.10% | 0.041% | 0.047% | 0.073 | 0.042 | 7.9e-02 | NA |
| MDD | Sjögren Syndrome, Possible (male) | 0.050 | 14,137 | 0.10% | 0.517% | 0.799% | 0.305 | 0.128 | 1.8e-02 | NA |
| MDD | Sjögren Syndrome, Probable (combined) | 1.000 | 99,216 | 0.10% | 0.042% | 0.057% | 0.081 | 0.051 | 1.1e-01 | NA |
| MDD | Sjögren Syndrome, Probable (female) | 0.200 | 36,975 | 0.10% | 0.028% | 0.035% | 0.064 | 0.054 | 2.4e-01 | NA |
| MDD | Sjögren Syndrome, Probable (male) | 0.500 | 67,956 | 0.10% | 0.375% | 0.605% | 0.265 | 0.158 | 9.3e-02 | NA |
| MDD | Systemic Lupus Erythematosus, Possible (combined) | 0.050 | 14,137 | 0.10% | 0.097% | 0.123% | 0.119 | 0.040 | 3.2e-03 | NA |
| MDD | Systemic Lupus Erythematosus, Possible (female) | 0.050 | 14,137 | 0.10% | 0.111% | 0.130% | 0.122 | 0.044 | 5.3e-03 | NA |
| MDD | Systemic Lupus Erythematosus, Possible (male) | 1.000 | 99,216 | 0.10% | 0.180% | 0.265% | 0.175 | 0.102 | 8.7e-02 | NA |
| MDD | Systemic Lupus Erythematosus, Probable (combined) | 0.050 | 14,137 | 0.10% | 0.170% | 0.231% | 0.163 | 0.053 | 2.1e-03 | NA |

| Base GWAS | Target Trait in UKB | Optimal p-value threshold ($P_T$) | No. SNPs used to construct PRS at ($P_T$) | Population Prevalence used to convert to liability $R^2$ | Unadjusted Variance explained (Observed $R^2$) | Adjusted Variance explained (Liability $R^2$) | Coefficient | Standard Error | P-value | Sex*PRS p-value |
|---|---|---|---|---|---|---|---|---|---|---|
| MDD | Systemic Lupus Erythematosus, Probable (female) | 0.050 | 14,137 | 0.10% | 0.190% | 0.242% | 0.166 | 0.057 | 3.5e-03 | NA |
| MDD | Systemic Lupus Erythematosus, Probable (male) | 0.500 | 67,956 | 0.10% | 0.154% | 0.243% | 0.168 | 0.141 | 2.3e-01 | NA |
| MDD | Type 1 Diabetes, Possible (combined) | 0.400 | 59,001 | 0.30% | 0.082% | 0.098% | 0.096 | 0.019 | 6.0e-07 | NA |
| MDD | Type 1 Diabetes, Possible (female) | 1.000 | 99,216 | 0.30% | 0.126% | 0.156% | 0.121 | 0.030 | 4.5e-05 | 0.18 |
| MDD | Type 1 Diabetes, Possible (male) | 0.300 | 48,852 | 0.30% | 0.066% | 0.075% | 0.084 | 0.025 | 8.7e-04 | NA |
| MDD | Type 1 Diabetes, Probable (combined) | 0.500 | 67,956 | 0.30% | 0.089% | 0.110% | 0.102 | 0.021 | 1.4e-06 | NA |
| MDD | Type 1 Diabetes, Probable (female) | 1.000 | 99,216 | 0.30% | 0.117% | 0.149% | 0.119 | 0.032 | 2.2e-04 | NA |
| MDD | Type 1 Diabetes, Probable (male) | 0.300 | 48,852 | 0.30% | 0.081% | 0.097% | 0.095 | 0.028 | 6.1e-04 | NA |

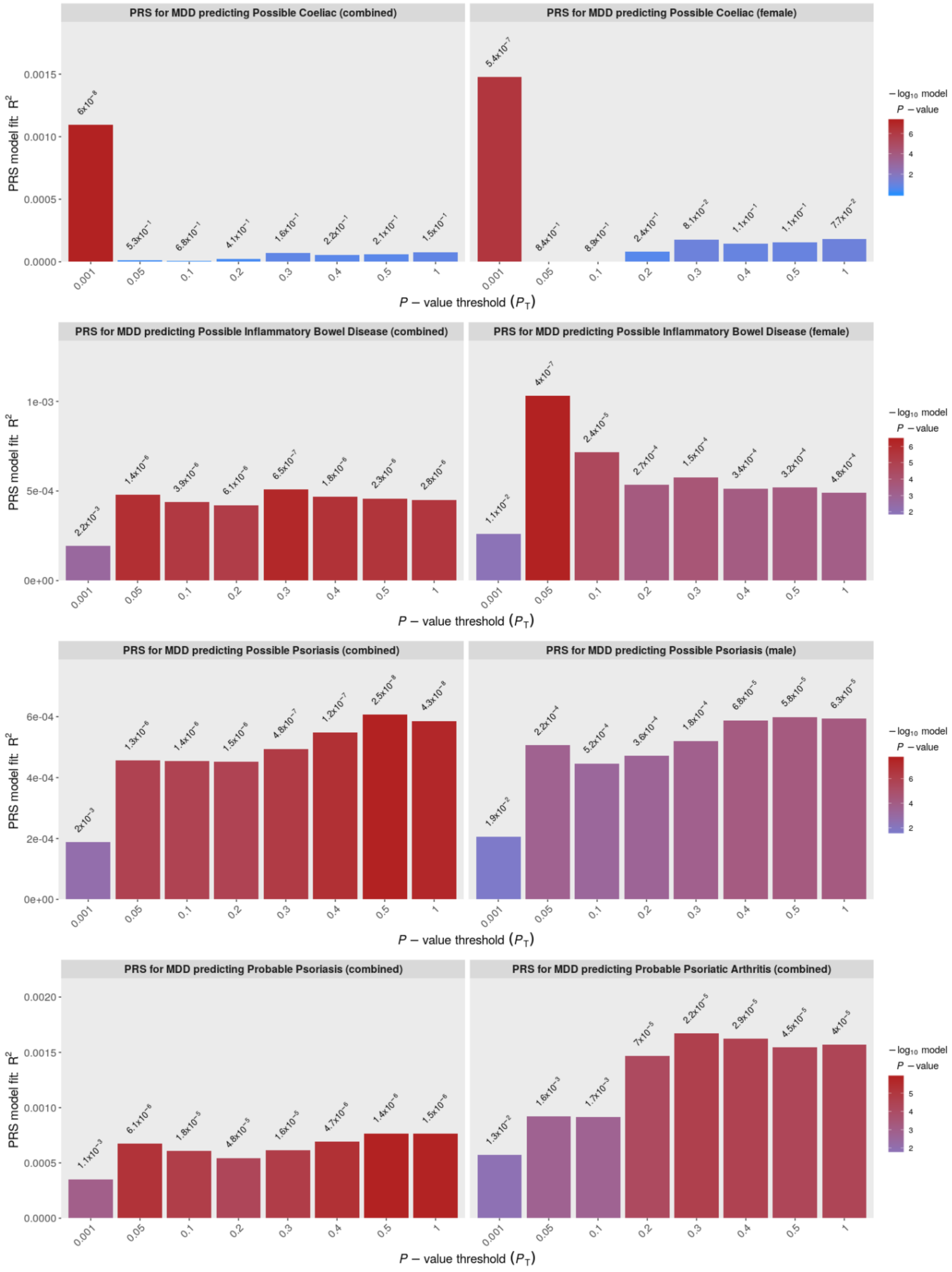
PRS for MDD predicting Possible Coeliac (combined)
PRS for MDD predicting Possible Coeliac (female)
PRS for MDD predicting Possible Inflammatory Bowel Disease (combined)
PRS for MDD predicting Possible Inflammatory Bowel Disease (female)
PRS for MDD predicting Possible Psoriasis (combined)
PRS for MDD predicting Possible Psoriasis (male)
PRS for MDD predicting Probable Psoriasis (combined)
PRS for MDD predicting Probable Psoriatic Arthritis (combined)
PRS model fit: $R^2$
$P$ – value threshold ($P_T$)
$-\log_{10}$ model $P$ – value
0.001
0.05
0.1
0.2
0.3
0.4
0.5
1

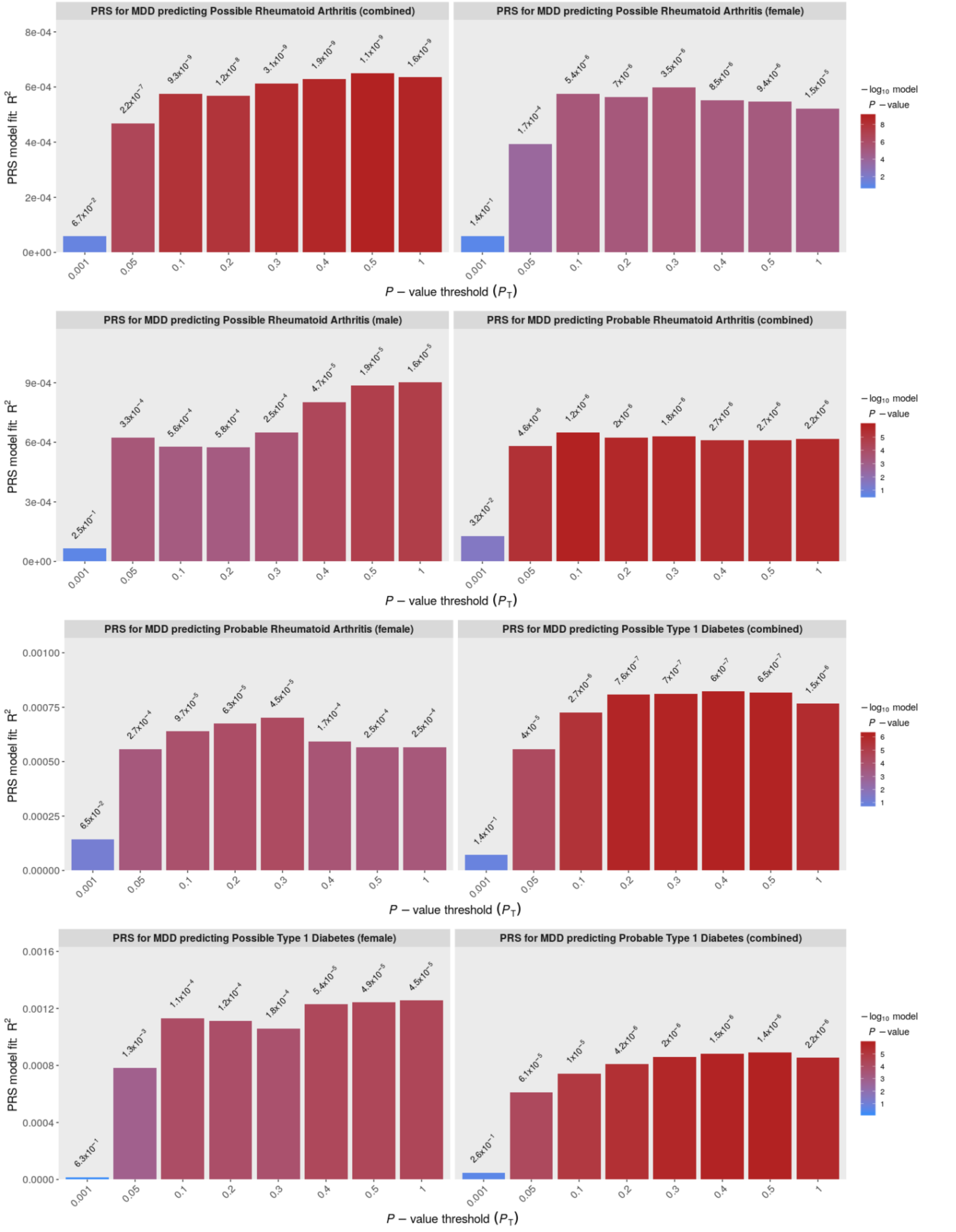

**Supplementary Figure 7**: Cross-trait PRS –associations between PRS for MDD and autoimmune case/control status (possible/probable) in men and women across eight $P_T$. Y-axis shows unadjusted $R^2$ (observed-scale). Bar charts are shown for significant results only.

### 3 Genetic correlations

**Supplementary Table 7:** Genetic correlations between autoimmune traits and ‘any’ and ‘stringent’ depression in the UKB. Associations with p-values < 4.1x10$^{-3}$, meeting Bonferroni correction, highlighted in red.

| | $r_G$ | 95% CIs | P-value | $r_G$ | 95% CIs | P-value |
|---|---|---|---|---|---|---|
| Autoimmune Disease | Any depression | | | Stringent depression | | |
| Coeliac Disease | 0.017 | -0.10 - 0.14 | 7.8e-01 | 0.048 | -0.10 - 0.19 | 5.2e-01 |
| Inflammatory Bowel Disease | 0.106 | 0.03 - 0.18 | 3.8e-03 | 0.157 | 0.07 - 0.24 | 3.0e-04 |
| Multiple Sclerosis | 0.302 | 0.06 - 0.55 | 1.6e-02 | 0.375 | 0.07 - 0.68 | 1.5e-02 |
| Psoriasis | 0.064 | -0.01 - 0.14 | 9.0e-02 | 0.161 | 0.06 - 0.26 | 1.1e-03 |
| Rheumatoid Arthritis | 0.004 | -0.06 - 0.07 | 9.0e-01 | 0.104 | 0.02 - 0.19 | 1.7e-02 |
| Systemic Lupus Erythematosus | -0.001 | -0.11 - 0.10 | 9.9e-01 | 0.126 | 0.00 - 0.25 | 5.1e-02 |
