## Supplementary Material for "Investigating pleiotropy between depression and autoimmune diseases using the UK Biobank"

Investigating pleiotropy between depression and autoimmune diseases using the UK Biobank, applying strict and minimal phenotyping, Supplementary Material

### Investigating pleiotropy between depression and autoimmune diseases using the UK Biobank, applying strict and minimal phenotyping, Supplementary Material

### 1 Definition of each autoimmune disease

#### 1.1 Pernicious Anemia

---

**Venn diagrams:** *Left:* probable cases as a subset of possible; *Right:* overlapping measures in the UK Biobank

**Matrix of ICD-10 primary and secondary dx - cells show number of individuals with each combination of dx**

|  | 0 primary dx | 1 primary dx | >1 primary dx |
| --- | --- | --- | --- |
| 0 secondary dx | 324,074^1^ | 24^2^ | 3^3^ |
| 1 secondary dx | 427^2^ | 3^3^ | 1^3^ |
| >1 secondary dx | 219^3^ | 3^3^ | 1^3^ |
|  |  |  |  |
| --- | --- | --- | --- |
| Note: |  |  |  |
| ^1^ Controls |  |  |  |
| ^2^ Possible cases |  |  |  |
| ^3^ Probable cases |  |  |  |

**ICD-10 codes used to construct matrix above. Columns show count of individuals who have received at least one dx**

| ICD-10 sub-codes | Primary dx | Secondary dx |
| --- | --- | --- |
| **D51 Vitamin B 12 deficiency anaemia** | | |
| D51.0 Vitamin B12 deficiency anaemia due to intrinsic factor deficiency | 35 | 654 |

**Count of self-reported autoimmune condition**

| Self-reported disorder | Number of people |
| --- | --- |
| Pernicious anaemia | 1,209 |

#### 1.2 Autoimmune Thyroid Disease

---

**Venn diagrams:** *Left:* probable cases as a subset of possible; *Right:* overlapping measures in the UK Biobank

**Matrix of ICD-10 primary and secondary dx - cells show number of individuals with each combination of dx**

|  | 0 primary dx | 1 primary dx | >1 primary dx |
| --- | --- | --- | --- |
| 0 secondary dx | 324,074^1^ | 147^2^ | 20^3^ |
| 1 secondary dx | 283^2^ | 25^3^ | 5^3^ |
| >1 secondary dx | 103^3^ | 11^3^ | 19^3^ |
|  |  |  |  |
| --- | --- | --- | --- |
| Note: |  |  |  |
| ^1^ Controls |  |  |  |
| ^2^ Possible cases |  |  |  |
| ^3^ Probable cases |  |  |  |

**ICD-10 codes used to construct matrix above. Columns show count of individuals who have received at least one dx**

| ICD-10 sub-codes | Primary dx | Secondary dx |
| --- | --- | --- |
| **E05 Thyrotoxicosis [hyperthyroidism]** | | |
| E05.0 Thyrotoxicosis with diffuse goitre | 169 | 297 |
| **E06 Thyroiditis** | | |
| E06.3 Autoimmune thyroiditis | 58 | 150 |

**Count of self-reported autoimmune condition**

| Self-reported disorder | Number of people |
| --- | --- |
| Thyroiditis | 200 |
| Grave’s disease | 100 |

**Count of self-reported prescription medications**

| Disease | Medication | Count |
| --- | --- | --- |
| Autoimmune Thyroid Disease (Grave’s and Thyroiditis) | levothyroxine sodium | 16,280 |
|  | thyroxine product | 4,605 |
|  | thyroxine sodium | 1,102 |
|  | carbimazole | 324 |
|  | sodium thyroxine | 183 |
|  | liothyronine | 95 |
|  | eltroxin 25micrograms tablet | 90 |
|  | propylthiouracil | 13 |
|  | sodium liothyronine | 9 |
|  | t3 - liothyronine | 6 |
|  | propylthiouracil product | 6 |
|  | neo-mercazole 5mg tablet | 4 |
|  | tertroxin 20mcg tablet | 4 |
|  | protirelin | 1 |

#### 1.3 Type 1 diabetes

---

**Venn diagrams:** *Left:* probable cases as a subset of possible; *Right:* overlapping measures in the UK Biobank

**Matrix of ICD-10 primary and secondary dx - cells show number of individuals with each combination of dx**

|  | 0 primary dx | 1 primary dx | >1 primary dx |
| --- | --- | --- | --- |
| 0 secondary dx | 324,074^1^ | 119^2^ | 37^3^ |
| 1 secondary dx | 1,029^2^ | 77^3^ | 34^3^ |
| >1 secondary dx | 987^3^ | 167^3^ | 198^3^ |
|  |  |  |  |
| --- | --- | --- | --- |
| Note: |  |  |  |
| ^1^ Controls |  |  |  |
| ^2^ Possible cases |  |  |  |
| ^3^ Probable cases |  |  |  |

**ICD-10 codes used to construct matrix above. Columns show count of individuals who have received at least one dx**

| ICD-10 sub-codes | Primary dx | Secondary dx |
| --- | --- | --- |
| **E10 Type 1 diabetes mellitus** | | |
| E10.0 With coma | 20 | 13 |
| E10.1 With ketoacidosis | 187 | 69 |
| E10.2 With renal complications | 10 | 71 |
| E10.3 With ophthalmic complications | 174 | 364 |
| E10.4 With neurological complications | 14 | 149 |
| E10.5 With peripheral circulatory complications | 56 | 57 |
| E10.6 With other specified complications | 16 | 27 |
| E10.7 With multiple complications | 0 | 8 |
| E10.8 With unspecified complications | 21 | 17 |
| E10.9 Without complications | 270 | 2,364 |

**Count of self-reported autoimmune condition**

| Self-reported disorder | Number of people |
| --- | --- |
| Type 1 diabetes | 364 |

**Count of self-reported prescription medications**

| Disease | Medication | Count |
| --- | --- | --- |
| Type 1 diabetes | insulin product | 3,925 |

#### 1.4 Multiple Sclerosis

---

**Venn diagrams:** *Left:* probable cases as a subset of possible; *Right:* overlapping measures in the UK Biobank

**Matrix of ICD-10 primary and secondary dx - cells show number of individuals with each combination of dx**

|  | 0 primary dx | 1 primary dx | >1 primary dx |
| --- | --- | --- | --- |
| 0 secondary dx | 324,074^1^ | 125^2^ | 97^3^ |
| 1 secondary dx | 292^2^ | 76^3^ | 43^3^ |
| >1 secondary dx | 325^3^ | 85^3^ | 209^3^ |
|  |  |  |  |
| --- | --- | --- | --- |
| Note: |  |  |  |
| ^1^ Controls |  |  |  |
| ^2^ Possible cases |  |  |  |
| ^3^ Probable cases |  |  |  |

**ICD-10 codes used to construct matrix above. Columns show count of individuals who have received at least one dx**

| ICD-10 sub-codes | Primary dx | Secondary dx |
| --- | --- | --- |
| **G35 Multiple sclerosis** | | |
| G35 Multiple sclerosis | 635 | 1,030 |

**Count of self-reported autoimmune condition**

| Self-reported disorder | Number of people |
| --- | --- |
| Multiple sclerosis | 1,426 |

**Count of self-reported prescription medications**

| Disease | Medication | Count |
| --- | --- | --- |
| Multiple Sclerosis | prednisolone | 2,310 |
|  | methotrexate | 2,096 |
|  | baclofen | 432 |
|  | mycophenolate | 174 |
|  | tacrolimus | 172 |
|  | prednisolone product | 139 |
|  | amantadine | 116 |
|  | ciclosporin | 50 |
|  | mtx - methotrexate | 36 |
|  | methylprednisolone | 19 |
|  | azt - azathioprine | 13 |
|  | myfortic 180mg gastro-resistant tablet | 13 |
|  | interferon beta-1a | 11 |
|  | glatiramer | 3 |
|  | interferon beta-1b | 3 |
|  | ciclosporin product | 1 |
|  | peginterferon alfa-2b | 1 |
|  | interferon beta-1b product | 0 |

#### 1.5 Myasthenia Gravis

---

**Venn diagrams:** *Left:* probable cases as a subset of possible; *Right:* overlapping measures in the UK Biobank

**Matrix of ICD-10 primary and secondary dx - cells show number of individuals with each combination of dx**

|  | 0 primary dx | 1 primary dx | >1 primary dx |
| --- | --- | --- | --- |
| 0 secondary dx | 324,074^1^ | 35^2^ | 7^3^ |
| 1 secondary dx | 32^2^ | 16^3^ | 11^3^ |
| >1 secondary dx | 44^3^ | 14^3^ | 22^3^ |
|  |  |  |  |
| --- | --- | --- | --- |
| Note: |  |  |  |
| ^1^ Controls |  |  |  |
| ^2^ Possible cases |  |  |  |
| ^3^ Probable cases |  |  |  |

**ICD-10 codes used to construct matrix above. Columns show count of individuals who have received at least one dx**

| ICD-10 sub-codes | Primary dx | Secondary dx |
| --- | --- | --- |
| **G70 Myasthenia gravis and other myoneural disorders** | | |
| G70.0 Myasthenia gravis | 105 | 139 |

**Count of self-reported autoimmune condition**

| Self-reported disorder | Number of people |
| --- | --- |
| Myasthenia gravis | 156 |

**Count of self-reported prescription medications**

| Disease | Medication | Count |
| --- | --- | --- |
| Myasthenia Gravis | prednisolone | 2,310 |
|  | methotrexate | 2,096 |
|  | mycophenolate | 174 |
|  | prednisolone product | 139 |
|  | ciclosporin | 50 |
|  | mtx - methotrexate | 36 |
|  | methylprednisolone | 19 |
|  | azt - azathioprine | 13 |
|  | myfortic 180mg gastro-resistant tablet | 13 |
|  | ciclosporin product | 1 |

#### 1.6 Coeliac

---

**Venn diagrams:** *Left:* probable cases as a subset of possible; *Right:* overlapping measures in the UK Biobank

**Matrix of ICD-10 primary and secondary dx - cells show number of individuals with each combination of dx**

|  | 0 primary dx | 1 primary dx | >1 primary dx |
| --- | --- | --- | --- |
| 0 secondary dx | 324,074^1^ | 400^2^ | 69^3^ |
| 1 secondary dx | 500^2^ | 148^3^ | 39^3^ |
| >1 secondary dx | 351^3^ | 134^3^ | 55^3^ |
|  |  |  |  |
| --- | --- | --- | --- |
| Note: |  |  |  |
| ^1^ Controls |  |  |  |
| ^2^ Possible cases |  |  |  |
| ^3^ Probable cases |  |  |  |

**ICD-10 codes used to construct matrix above. Columns show count of individuals who have received at least one dx**

| ICD-10 sub-codes | Primary dx | Secondary dx |
| --- | --- | --- |
| **K90 Intestinal malabsorption** | | |
| K90.0 Coeliac disease | 845 | 1,227 |

**Count of self-reported autoimmune condition**

| Self-reported disorder | Number of people |
| --- | --- |
| Malabsorption/coeliac disease | 1,704 |

#### 1.7 Inflammatory Bowel Disease

---

**Venn diagrams:** *Left:* probable cases as a subset of possible; *Right:* overlapping measures in the UK Biobank

**Matrix of ICD-10 primary and secondary dx - cells show number of individuals with each combination of dx**

|  | 0 primary dx | 1 primary dx | >1 primary dx |
| --- | --- | --- | --- |
| 0 secondary dx | 324,074^1^ | 988^2^ | 517^3^ |
| 1 secondary dx | 597^2^ | 318^3^ | 382^3^ |
| >1 secondary dx | 328^3^ | 283^3^ | 813^3^ |
|  |  |  |  |
| --- | --- | --- | --- |
| Note: |  |  |  |
| ^1^ Controls |  |  |  |
| ^2^ Possible cases |  |  |  |
| ^3^ Probable cases |  |  |  |

**ICD-10 codes used to construct matrix above. Columns show count of individuals who have received at least one dx**

| ICD-10 sub-codes | Primary dx | Secondary dx |
| --- | --- | --- |
| **K50 Crohn disease [regional enteritis]** | | |
| K50.0 Crohn’s disease of small intestine | 264 | 166 |
| K50.1 Crohn’s disease of large intestine | 356 | 165 |
| K50.8 Other Crohn’s disease | 111 | 58 |
| K50.9 Crohn’s disease, unspecified | 814 | 1,013 |
| **K51 Ulcerative colitis** | | |
| K51.0 Ulcerative (chronic) enterocolitis | 101 | 34 |
| K51.1 Ulcerative (chronic) ileocolitis | 20 | 7 |
| K51.2 Ulcerative (chronic) proctitis | 328 | 81 |
| K51.3 Ulcerative (chronic) rectosigmoiditis | 174 | 34 |
| K51.4 Pseudopolyposis of colon | 99 | 99 |
| K51.5 Mucosal proctocolitis | 69 | 13 |
| K51.8 Other ulcerative colitis | 172 | 62 |
| K51.9 Ulcerative colitis, unspecified | 2,027 | 1,573 |

**Count of self-reported autoimmune condition**

| Self-reported disorder | Number of people |
| --- | --- |
| Crohns disease | 1,186 |
| Ulcerative colitis | 2,114 |

**Count of self-reported prescription medications**

| Disease | Medication | Count |
| --- | --- | --- |
| Inflammatory Bowel Disease | prednisolone | 2,310 |
|  | methotrexate | 2,096 |
|  | mesalazine | 570 |
|  | asacol 400mg e/c tablet | 563 |
|  | pentasa sr 250mg m/r tablet | 263 |
|  | asacol mr 400mg e/c tablet | 155 |
|  | prednisolone product | 139 |
|  | balsalazide disodium | 118 |
|  | humira 40mg injection solution 0.8ml prefilled syringe | 104 |
|  | olsalazine | 47 |
|  | adalimumab | 43 |
|  | mtx - methotrexate | 36 |
|  | methylprednisolone | 19 |
|  | azt - azathioprine | 13 |
|  | 5asa - mesalazine | 7 |

#### 1.8 Psoriasis

---

**Venn diagrams:** *Left:* probable cases as a subset of possible; *Right:* overlapping measures in the UK Biobank

**Matrix of ICD-10 primary and secondary dx - cells show number of individuals with each combination of dx**

|  | 0 primary dx | 1 primary dx | >1 primary dx |
| --- | --- | --- | --- |
| 0 secondary dx | 324,074^1^ | 90^2^ | 92^3^ |
| 1 secondary dx | 932^2^ | 31^3^ | 18^3^ |
| >1 secondary dx | 314^3^ | 14^3^ | 39^3^ |
|  |  |  |  |
| --- | --- | --- | --- |
| Note: |  |  |  |
| ^1^ Controls |  |  |  |
| ^2^ Possible cases |  |  |  |
| ^3^ Probable cases |  |  |  |

**ICD-10 codes used to construct matrix above. Columns show count of individuals who have received at least one dx**

| ICD-10 sub-codes | Primary dx | Secondary dx |
| --- | --- | --- |
| **L40 Psoriasis** | | |
| L40.0 Psoriasis vulgaris | 86 | 51 |
| L40.1 Generalised pustular psoriasis | 9 | 7 |
| L40.3 Pustulosis palmaris et plantaris | 8 | 8 |
| L40.4 Guttate psoriasis | 5 | 8 |
| L40.8 Other psoriasis | 16 | 17 |
| L40.9 Psoriasis, unspecified | 213 | 1,295 |

**Count of self-reported autoimmune condition**

| Self-reported disorder | Number of people |
| --- | --- |
| Psoriasis | 4,664 |

**Count of self-reported prescription medications**

| Disease | Medication | Count |
| --- | --- | --- |
| Psoriasis | beclometasone | 3,582 |
|  | prednisolone | 2,310 |
|  | methotrexate | 2,096 |
|  | beclomethasone | 1,594 |
|  | vitamin d product | 1,463 |
|  | betnovate cream | 1,384 |
|  | mometasone | 816 |
|  | hydrocortisone | 731 |
|  | diprobase cream | 644 |
|  | dermovate cream | 642 |
|  | betamethasone | 634 |
|  | dovobet ointment | 476 |
|  | eumovate cream | 427 |
|  | aqueous cream bp | 330 |
|  | hydrocortisone product | 271 |
|  | e45 cream | 269 |
|  | elocon cream | 263 |
|  | daktacort cream | 263 |
|  | trimovate ointment | 257 |
|  | prednisone | 251 |
|  | epaderm ointment | 243 |
|  | calcipotriol | 239 |
|  | dovonex 50micrograms/g cream | 222 |
|  | doublebase gel | 209 |
|  | fucibet cream | 204 |
|  | diprosalic ointment | 187 |
|  | mycophenolate | 174 |
|  | prednisolone product | 139 |
|  | dermol 500 lotion | 122 |
|  | clobetasone | 120 |
|  | humira 40mg injection solution 0.8ml prefilled syringe | 104 |
|  | synalar 1:10 cream | 101 |
|  | capasal shampoo | 100 |
|  | dovonex 50micrograms/g ointment | 97 |
|  | polytar liquid | 96 |
|  | clobetasol | 95 |
|  | mometasone furoate 0.1% ointment | 93 |
|  | clotrimazole | 91 |
|  | cetraben emollient cream | 88 |
|  | dovonex scalp solution | 81 |
|  | cetraben cream | 80 |
|  | dermol cream | 75 |
|  | oilatum emollient bath additive | 74 |
|  | diprobase ointment | 72 |
|  | locoid 0.1% cream | 69 |
|  | protopic 0.03% ointment | 67 |
|  | timodine cream | 67 |
|  | exorex lotion | 66 |
|  | hydromol cream | 66 |
|  | ketoconazole 2% shampoo | 65 |
|  | fucidin cream | 65 |
|  | mometasone furoate 0.1% lotion | 63 |
|  | cortisone | 62 |
|  | cyclosporin | 61 |
|  | neoral 10mg capsule | 59 |
|  | emulsifying ointment bp | 57 |
|  | betamethasone+calcipotriol | 55 |
|  | betacap scalp application | 53 |
|  | ciclosporin | 50 |
|  | neotigason 10mg capsule | 47 |
|  | nizoral 20mg/ml shampoo | 47 |
|  | cocois ointment | 46 |
|  | oilatum bath formula liquid bath additive | 44 |
|  | adalimumab | 43 |
|  | acitretin | 42 |
|  | calcitriol | 42 |
|  | cortisone product | 41 |
|  | alphosyl cream | 39 |
|  | balneum bath oil | 39 |
|  | fusidic acid | 39 |
|  | dermol 200 shower emollient | 38 |
|  | unguentum m cream | 37 |
|  | mtx - methotrexate | 36 |
|  | alphosyl hc cream | 35 |
|  | ketoconazole | 33 |
|  | metosyn 0.05% cream | 33 |
|  | liquid paraffin product | 31 |
|  | canesten hc cream | 31 |
|  | hydrocortisone+miconazole | 31 |
|  | tioconazole | 30 |
|  | coal tar product | 29 |
|  | alphaderm cream | 29 |
|  | cocois scalp ointment | 28 |
|  | diprosone cream | 28 |
|  | hydrocortisone+clotrimazole | 28 |
|  | emollient product | 27 |
|  | alphosyl shampoo | 26 |
|  | lotriderm cream | 26 |
|  | hydrocortistab 1% cream | 25 |
|  | liquid paraffin | 23 |
|  | salicylic acid product | 23 |
|  | curatoderm 4micrograms/g ointment | 22 |
|  | eurax cream | 22 |
|  | balneum plus cream | 21 |
|  | fucidin h cream | 21 |
|  | silkis 3micrograms/g ointment | 20 |
|  | calcitriol product | 20 |
|  | liquid paraffin+white soft paraffin 50%/50% ointment | 20 |
|  | balneum plus bath oil | 20 |
|  | nerisone 0.1% cream | 20 |
|  | polytar af liquid | 18 |
|  | clobetasone butyrate+neomycin sulphate | 18 |
|  | cutivate 0.05% cream | 17 |
|  | unguentum merck cream | 17 |
|  | nizoral 2% cream | 16 |
|  | hydrocortisyl 1% cream | 16 |
|  | cya - cyclosporin | 16 |
|  | elidel 1% cream | 16 |
|  | alphosyl 2 in 1 shampoo | 15 |
|  | clobetasol propionate+neomycin sulphate+nystatin | 15 |
|  | hydromol emollient bath additive | 15 |
|  | fucidin ointment | 14 |
|  | e45 lotion | 13 |
|  | tacrolimus monohydrate 0.03% ointment | 13 |
|  | polytar plus liquid | 12 |
|  | polytar emollient bath additive | 12 |
|  | aveeno lotion | 12 |
|  | dithrocream 0.1% cream | 11 |
|  | urea 10% cream | 11 |
|  | hc - hydrocortisone | 11 |
|  | eucerin 10% cream | 11 |
|  | tacalcitol | 10 |
|  | paraffin-white soft | 10 |
|  | haelan 0.0125% cream | 10 |
|  | salicylic acid | 9 |
|  | paraffin liquid | 9 |
|  | diprobath bath additive | 9 |
|  | e45 emollient bath oil | 9 |
|  | betamethasone+salicylic acid | 8 |
|  | emulsiderm emollient emulsion | 7 |
|  | selsun shampoo | 7 |
|  | calmurid hc cream | 7 |
|  | hydrocortisone+fusidic acid | 7 |
|  | eurax hc cream | 7 |
|  | dithranol | 6 |
|  | coal tar 4.3% shampoo | 6 |
|  | sebco ointment | 6 |
|  | psoriderm cream | 6 |
|  | bettamousse 0.12% foam | 6 |
|  | neutrogena dermatological cream | 5 |
|  | modrasone 0.05% cream | 5 |
|  | ung emuls - ungentum emulsificans | 5 |
|  | dermol cream 500g | 5 |
|  | mycophenolic acid 180mg gastro-resistant tablet | 5 |
|  | coal tar+salicylic acid ointment bp | 4 |
|  | t/gel shampoo | 4 |
|  | alphosyl lotion | 4 |
|  | dermacare cream 100ml | 4 |
|  | dithranol 0.2% ointment bp | 3 |
|  | dithrocream 2% cream | 3 |
|  | tazarotene | 3 |
|  | dithrocream forte 0.5% cream | 3 |
|  | dithrocream hp 1% cream | 3 |
|  | coal tar extract+hydrocortisone 3%/0.25% cream | 3 |
|  | coal tar extract 2% shampoo | 3 |
|  | efalizumab | 3 |
|  | clobetasone butyrate+oxytetracycline+nystatin | 3 |
|  | dithrocream 0.25% cream | 2 |
|  | psoriderm shampoo | 2 |
|  | hydrous ointment bp | 2 |
|  | dithranol+salicylic acid 0.25%/1.6% scalp gel | 2 |
|  | urea 10% lotion | 2 |
|  | oilatum bath formula liquid bath additive 150ml | 2 |
|  | zorac 0.05% aqueous gel | 1 |
|  | coal tar 40% bath emulsion | 1 |
|  | coal tar extract+allantoin 5%/2% cream | 1 |
|  | psoriderm bath emulsion | 1 |
|  | coal tar+lecithin 6/0.4% cream | 1 |
|  | strong coal tar solution+pine tar 5/5% gel | 1 |
|  | coal tar extract+allantoin 5%/2% lotion | 1 |
|  | psorin scalp gel | 1 |
|  | urea+lauromacrogols 5%/3% cream | 1 |
|  | dermacare cream 150ml | 1 |
|  | cortacream 1% band | 0 |
|  | methyl salicylate ointment bp | 0 |
|  | meted shampoo | 0 |

#### 1.9 Ankylosing Spondylitis

---

**Venn diagrams:** *Left:* probable cases as a subset of possible; *Right:* overlapping measures in the UK Biobank

**Matrix of ICD-10 primary and secondary dx - cells show number of individuals with each combination of dx**

|  | 0 primary dx | 1 primary dx | >1 primary dx |
| --- | --- | --- | --- |
| 0 secondary dx | 324,074^1^ | 36^2^ | 25^3^ |
| 1 secondary dx | 266^2^ | 13^3^ | 12^3^ |
| >1 secondary dx | 147^3^ | 27^3^ | 40^3^ |
|  |  |  |  |
| --- | --- | --- | --- |
| Note: |  |  |  |
| ^1^ Controls |  |  |  |
| ^2^ Possible cases |  |  |  |
| ^3^ Probable cases |  |  |  |

**ICD-10 codes used to construct matrix above. Columns show count of individuals who have received at least one dx**

| ICD-10 sub-codes | Primary dx | Secondary dx |
| --- | --- | --- |
| **M45 Ankylosing spondylitis** | | |
| M45 Ankylosing spondylitis | 101 | 303 |
| M45.X0 Ankylosing spondylitis (Multiple sites in spine) | 40 | 35 |
| M45.X1 Ankylosing spondylitis (Occipito-atlanto-axial region) | 0 | 4 |
| M45.X2 Ankylosing spondylitis (Cervical region) | 9 | 29 |
| M45.X4 Ankylosing spondylitis (Thoracic region) | 0 | 1 |
| M45.X6 Ankylosing spondylitis (Lumbar region) | 7 | 6 |
| M45.X7 Ankylosing spondylitis (Lumbosacral region) | 3 | 4 |
| M45.X8 Ankylosing spondylitis (Sacral and sacrococcygeal region) | 0 | 2 |
| M45.X9 Ankylosing spondylitis (Site unspecified) | 50 | 250 |

**Count of self-reported autoimmune condition**

| Self-reported disorder | Number of people |
| --- | --- |
| Ankylosing spondylitis | 1,118 |

**Count of self-reported prescription medications**

| Disease | Medication | Count |
| --- | --- | --- |
| Ankylosing Spondylitis | azathioprine | 841 |
|  | humira 40mg injection solution 0.8ml prefilled syringe | 104 |
|  | adalimumab | 43 |

#### 1.10 Polymyalgia Rheumatica/Giant Cell Arteritis

---

**Venn diagrams:** *Left:* probable cases as a subset of possible; *Right:* overlapping measures in the UK Biobank

**Matrix of ICD-10 primary and secondary dx - cells show number of individuals with each combination of dx**

|  | 0 primary dx | 1 primary dx | >1 primary dx |
| --- | --- | --- | --- |
| 0 secondary dx | 324,074^1^ | 46^2^ | 6^3^ |
| 1 secondary dx | 573^2^ | 19^3^ | 3^3^ |
| >1 secondary dx | 384^3^ | 24^3^ | 7^3^ |
|  |  |  |  |
| --- | --- | --- | --- |
| Note: |  |  |  |
| ^1^ Controls |  |  |  |
| ^2^ Possible cases |  |  |  |
| ^3^ Probable cases |  |  |  |

**ICD-10 codes used to construct matrix above. Columns show count of individuals who have received at least one dx**

| ICD-10 sub-codes | Primary dx | Secondary dx |
| --- | --- | --- |
| **M31 Other necrotizing vasculopathies** | | |
| M31.5 Giant cell arteritis with polymyalgia rheumatica | 11 | 33 |
| **M35 Other systemic involvement of connective tissue** | | |
| M35.3 Polymyalgia rheumatica | 96 | 1,002 |

**Count of self-reported autoimmune condition**

| Self-reported disorder | Number of people |
| --- | --- |
| Polymyalgia rheumatica | 871 |

**Count of self-reported prescription medications**

| Disease | Medication | Count |
| --- | --- | --- |
| Polymyalgia Rheumatica/Giant Cell Arteritis | prednisolone | 2,310 |
|  | methotrexate | 2,096 |
|  | prednisolone product | 139 |
|  | mtx - methotrexate | 36 |
|  | methylprednisolone | 19 |
|  | azt - azathioprine | 13 |

#### 1.11 Psoriatic Arthritis

---

**Venn diagrams:** *Left:* probable cases as a subset of possible; *Right:* overlapping measures in the UK Biobank

**Matrix of ICD-10 primary and secondary dx - cells show number of individuals with each combination of dx**

|  | 0 primary dx | 1 primary dx | >1 primary dx |
| --- | --- | --- | --- |
| 0 secondary dx | 324,074^1^ | 11^2^ | 8^3^ |
| 1 secondary dx | 21^2^ | 69^3^ | 0^3^ |
| >1 secondary dx | 373^3^ | 68^3^ | 116^3^ |
|  |  |  |  |
| --- | --- | --- | --- |
| Note: |  |  |  |
| ^1^ Controls |  |  |  |
| ^2^ Possible cases |  |  |  |
| ^3^ Probable cases |  |  |  |

**ICD-10 codes used to construct matrix above. Columns show count of individuals who have received at least one dx**

| ICD-10 sub-codes | Primary dx | Secondary dx |
| --- | --- | --- |
| **L40 Psoriasis** | | |
| L40.5 Arthropathic psoriasis | 258 | 554 |
| **M07 Psoriatic and enteropathic arthropathies** | | |
| M07.3 Other psoriatic arthropathies | 7 | 354 |
| M07.30 Other psoriatic arthropathies (Multiple sites) | 6 | 139 |
| M07.31 Other psoriatic arthropathies (Shoulder region) | 0 | 7 |
| M07.32 Other psoriatic arthropathies (Upper arm) | 0 | 1 |
| M07.33 Other psoriatic arthropathies (Forearm) | 0 | 5 |
| M07.34 Other psoriatic arthropathies (Hand) | 0 | 19 |
| M07.35 Other psoriatic arthropathies (Pelvic region and thigh) | 1 | 15 |
| M07.36 Other psoriatic arthropathies (Lower leg) | 5 | 29 |
| M07.37 Other psoriatic arthropathies (Ankle and foot) | 2 | 21 |
| M07.38 Other psoriatic arthropathies (Other) | 0 | 7 |
| M07.39 Other psoriatic arthropathies (Site unspecified) | 7 | 260 |

**Count of self-reported autoimmune condition**

| Self-reported disorder | Number of people |
| --- | --- |
| Psoriatic arthropathy | 766 |

**Count of self-reported prescription medications**

| Disease | Medication | Count |
| --- | --- | --- |
| Psoriatic Arthritis | prednisolone | 2,310 |
|  | methotrexate | 2,096 |
|  | azathioprine | 841 |
|  | leflunomide | 158 |
|  | prednisolone product | 139 |
|  | humira 40mg injection solution 0.8ml prefilled syringe | 104 |
|  | adalimumab | 43 |
|  | mtx - methotrexate | 36 |
|  | methylprednisolone | 19 |
|  | arava 10mg tablet | 9 |
|  | arava 20mg tablet | 6 |

#### 1.12 Rheumatoid Arthritis

---

**Venn diagrams:** *Left:* probable cases as a subset of possible; *Right:* overlapping measures in the UK Biobank

**Matrix of ICD-10 primary and secondary dx - cells show number of individuals with each combination of dx**

|  | 0 primary dx | 1 primary dx | >1 primary dx |
| --- | --- | --- | --- |
| 0 secondary dx | 324,074^1^ | 376^2^ | 184^3^ |
| 1 secondary dx | 1,425^2^ | 121^3^ | 166^3^ |
| >1 secondary dx | 977^3^ | 254^3^ | 576^3^ |
|  |  |  |  |
| --- | --- | --- | --- |
| Note: |  |  |  |
| ^1^ Controls |  |  |  |
| ^2^ Possible cases |  |  |  |
| ^3^ Probable cases |  |  |  |

**ICD-10 codes used to construct matrix above. Columns show count of individuals who have received at least one dx**

| ICD-10 sub-codes | Primary dx | Secondary dx |
| --- | --- | --- |
| **M05 Seropositive rheumatoid arthritis** | | |
| M05.0 Felty’s syndrome | 1 | 4 |
| M05.00 Felty’s syndrome (Multiple sites) | 2 | 4 |
| M05.09 Felty’s syndrome (Site unspecified) | 1 | 4 |
| M05.1 Rheumatoid lung disease | 9 | 14 |
| M05.10 Rheumatoid lung disease (Multiple sites) | 3 | 3 |
| M05.19 Rheumatoid lung disease (Site unspecified) | 3 | 7 |
| M05.2 Rheumatoid vasculitis | 8 | 6 |
| M05.20 Rheumatoid vasculitis (Multiple sites) | 5 | 3 |
| M05.26 Rheumatoid vasculitis (Lower leg) | 1 | 0 |
| M05.28 Rheumatoid vasculitis (Other) | 1 | 0 |
| M05.29 Rheumatoid vasculitis (Site unspecified) | 4 | 5 |
| M05.3 Rheumatoid arthritis with involvement of other organs and systems | 1 | 2 |
| M05.30 Rheumatoid arthritis with involvement of other organs and systems (Multiple sites) | 1 | 1 |
| M05.38 Rheumatoid arthritis with involvement of other organs and systems (Other) | 0 | 1 |
| M05.8 Other seropositive rheumatoid arthritis | 12 | 6 |
| M05.80 Other seropositive rheumatoid arthritis (Multiple sites) | 8 | 4 |
| M05.82 Other seropositive rheumatoid arthritis (Upper arm) | 1 | 0 |
| M05.83 Other seropositive rheumatoid arthritis (Forearm) | 3 | 0 |
| M05.84 Other seropositive rheumatoid arthritis (Hand) | 2 | 1 |
| M05.86 Other seropositive rheumatoid arthritis (Lower leg) | 1 | 0 |
| M05.87 Other seropositive rheumatoid arthritis (Ankle and foot) | 1 | 0 |
| M05.88 Other seropositive rheumatoid arthritis (Other) | 1 | 0 |
| M05.89 Other seropositive rheumatoid arthritis (Site unspecified) | 5 | 2 |
| M05.9 Seropositive rheumatoid arthritis, unspecified | 90 | 107 |
| M05.90 Seropositive rheumatoid arthritis, unspecified (Multiple sites) | 164 | 101 |
| M05.91 Seropositive rheumatoid arthritis, unspecified (Shoulder region) | 4 | 3 |
| M05.92 Seropositive rheumatoid arthritis, unspecified (Upper arm) | 8 | 2 |
| M05.93 Seropositive rheumatoid arthritis, unspecified (Forearm) | 7 | 3 |
| M05.94 Seropositive rheumatoid arthritis, unspecified (Hand) | 11 | 5 |
| M05.95 Seropositive rheumatoid arthritis, unspecified (Pelvic region and thigh) | 7 | 3 |
| M05.96 Seropositive rheumatoid arthritis, unspecified (Lower leg) | 17 | 3 |
| M05.97 Seropositive rheumatoid arthritis, unspecified (Ankle and foot) | 19 | 4 |
| M05.98 Seropositive rheumatoid arthritis, unspecified (Other) | 2 | 1 |
| M05.99 Seropositive rheumatoid arthritis, unspecified (Site unspecified) | 68 | 81 |
| **M06 Other rheumatoid arthritis** | | |
| M06.0 Seronegative rheumatoid arthritis | 57 | 115 |
| M06.00 Seronegative rheumatoid arthritis (Multiple sites) | 105 | 94 |
| M06.01 Seronegative rheumatoid arthritis (Shoulder region) | 7 | 1 |
| M06.02 Seronegative rheumatoid arthritis (Upper arm) | 4 | 1 |
| M06.03 Seronegative rheumatoid arthritis (Forearm) | 4 | 1 |
| M06.04 Seronegative rheumatoid arthritis (Hand) | 3 | 5 |
| M06.05 Seronegative rheumatoid arthritis (Pelvic region and thigh) | 4 | 2 |
| M06.06 Seronegative rheumatoid arthritis (Lower leg) | 16 | 6 |
| M06.07 Seronegative rheumatoid arthritis (Ankle and foot) | 10 | 2 |
| M06.08 Seronegative rheumatoid arthritis (Other) | 1 | 0 |
| M06.09 Seronegative rheumatoid arthritis (Site unspecified) | 33 | 104 |
| M06.1 Adult-onset Still’s disease | 6 | 4 |
| M06.10 Adult-onset Still’s disease (Multiple sites) | 2 | 3 |
| M06.16 Adult-onset Still’s disease (Lower leg) | 1 | 0 |
| M06.19 Adult-onset Still’s disease (Site unspecified) | 3 | 3 |
| M06.21 Rheumatoid bursitis (Shoulder region) | 1 | 0 |
| M06.22 Rheumatoid bursitis (Upper arm) | 1 | 0 |
| M06.27 Rheumatoid bursitis (Ankle and foot) | 1 | 0 |
| M06.3 Rheumatoid nodule | 19 | 3 |
| M06.30 Rheumatoid nodule (Multiple sites) | 8 | 4 |
| M06.32 Rheumatoid nodule (Upper arm) | 20 | 5 |
| M06.33 Rheumatoid nodule (Forearm) | 5 | 3 |
| M06.34 Rheumatoid nodule (Hand) | 48 | 13 |
| M06.36 Rheumatoid nodule (Lower leg) | 2 | 2 |
| M06.37 Rheumatoid nodule (Ankle and foot) | 22 | 10 |
| M06.39 Rheumatoid nodule (Site unspecified) | 2 | 0 |
| M06.4 Inflammatory polyarthropathy | 23 | 19 |
| M06.40 Inflammatory polyarthropathy (Multiple sites) | 32 | 19 |
| M06.41 Inflammatory polyarthropathy (Shoulder region) | 2 | 1 |
| M06.43 Inflammatory polyarthropathy (Forearm) | 2 | 0 |
| M06.44 Inflammatory polyarthropathy (Hand) | 2 | 0 |
| M06.45 Inflammatory polyarthropathy (Pelvic region and thigh) | 1 | 1 |
| M06.46 Inflammatory polyarthropathy (Lower leg) | 9 | 2 |
| M06.47 Inflammatory polyarthropathy (Ankle and foot) | 2 | 0 |
| M06.49 Inflammatory polyarthropathy (Site unspecified) | 40 | 19 |
| M06.8 Other specified rheumatoid arthritis | 10 | 3 |
| M06.80 Other specified rheumatoid arthritis (Multiple sites) | 3 | 6 |
| M06.81 Other specified rheumatoid arthritis (Shoulder region) | 2 | 0 |
| M06.82 Other specified rheumatoid arthritis (Upper arm) | 1 | 1 |
| M06.84 Other specified rheumatoid arthritis (Hand) | 0 | 1 |
| M06.85 Other specified rheumatoid arthritis (Pelvic region and thigh) | 1 | 0 |
| M06.86 Other specified rheumatoid arthritis (Lower leg) | 1 | 0 |
| M06.87 Other specified rheumatoid arthritis (Ankle and foot) | 2 | 0 |
| M06.88 Other specified rheumatoid arthritis (Other) | 5 | 0 |
| M06.89 Other specified rheumatoid arthritis (Site unspecified) | 6 | 4 |
| M06.9 Rheumatoid arthritis, unspecified | 513 | 1,682 |
| M06.90 Rheumatoid arthritis, unspecified (Multiple sites) | 449 | 697 |
| M06.91 Rheumatoid arthritis, unspecified (Shoulder region) | 77 | 29 |
| M06.92 Rheumatoid arthritis, unspecified (Upper arm) | 42 | 4 |
| M06.93 Rheumatoid arthritis, unspecified (Forearm) | 41 | 25 |
| M06.94 Rheumatoid arthritis, unspecified (Hand) | 116 | 89 |
| M06.95 Rheumatoid arthritis, unspecified (Pelvic region and thigh) | 55 | 15 |
| M06.96 Rheumatoid arthritis, unspecified (Lower leg) | 137 | 65 |
| M06.97 Rheumatoid arthritis, unspecified (Ankle and foot) | 114 | 54 |
| M06.98 Rheumatoid arthritis, unspecified (Other) | 13 | 19 |
| M06.99 Rheumatoid arthritis, unspecified (Site unspecified) | 475 | 1,868 |

**Count of self-reported autoimmune condition**

| Self-reported disorder | Number of people |
| --- | --- |
| Rheumatoid arthritis | 4,364 |

**Count of self-reported prescription medications**

| Disease | Medication | Count |
| --- | --- | --- |
| Rheumatoid Arthritis | prednisolone | 2,310 |
|  | methotrexate | 2,096 |
|  | azathioprine | 841 |
|  | hydroxychloroquine | 590 |
|  | leflunomide | 158 |
|  | prednisolone product | 139 |
|  | humira 40mg injection solution 0.8ml prefilled syringe | 104 |
|  | adalimumab | 43 |
|  | mtx - methotrexate | 36 |
|  | methylprednisolone | 19 |
|  | azt - azathioprine | 13 |
|  | arava 10mg tablet | 9 |
|  | arava 20mg tablet | 6 |
|  | gold product | 6 |
|  | sodium aurothiomalate | 4 |
|  | gold sodium thiomalate | 2 |

#### 1.13 Sjögren Syndrome

---

**Venn diagrams:** *Left:* probable cases as a subset of possible; *Right:* overlapping measures in the UK Biobank

**Matrix of ICD-10 primary and secondary dx - cells show number of individuals with each combination of dx**

|  | 0 primary dx | 1 primary dx | >1 primary dx |
| --- | --- | --- | --- |
| 0 secondary dx | 324,074^1^ | 22^2^ | 4^3^ |
| 1 secondary dx | 186^2^ | 11^3^ | 8^3^ |
| >1 secondary dx | 204^3^ | 19^3^ | 15^3^ |
|  |  |  |  |
| --- | --- | --- | --- |
| Note: |  |  |  |
| ^1^ Controls |  |  |  |
| ^2^ Possible cases |  |  |  |
| ^3^ Probable cases |  |  |  |

**ICD-10 codes used to construct matrix above. Columns show count of individuals who have received at least one dx**

| ICD-10 sub-codes | Primary dx | Secondary dx |
| --- | --- | --- |
| **M35 Other systemic involvement of connective tissue** | | |
| M35.0 Sicca syndrome [Sjogren] | 79 | 443 |

**Count of self-reported autoimmune condition**

| Self-reported disorder | Number of people |
| --- | --- |
| Sjogren’s syndrome/sicca syndrome | 373 |

**Count of self-reported prescription medications**

| Disease | Medication | Count |
| --- | --- | --- |
| Sjögren Syndrome | prednisolone | 2,310 |
|  | methotrexate | 2,096 |
|  | hydroxychloroquine | 590 |
|  | prednisolone product | 139 |
|  | mtx - methotrexate | 36 |
|  | methylprednisolone | 19 |
|  | azt - azathioprine | 13 |

#### 1.14 Systemic Lupus Erythematosus

---

**Venn diagrams:** *Left:* probable cases as a subset of possible; *Right:* overlapping measures in the UK Biobank

**Matrix of ICD-10 primary and secondary dx - cells show number of individuals with each combination of dx**

|  | 0 primary dx | 1 primary dx | >1 primary dx |
| --- | --- | --- | --- |
| 0 secondary dx | 324,074^1^ | 43^2^ | 11^3^ |
| 1 secondary dx | 144^2^ | 8^3^ | 6^3^ |
| >1 secondary dx | 128^3^ | 27^3^ | 34^3^ |
|  |  |  |  |
| --- | --- | --- | --- |
| Note: |  |  |  |
| ^1^ Controls |  |  |  |
| ^2^ Possible cases |  |  |  |
| ^3^ Probable cases |  |  |  |

**ICD-10 codes used to construct matrix above. Columns show count of individuals who have received at least one dx**

| ICD-10 sub-codes | Primary dx | Secondary dx |
| --- | --- | --- |
| **L93 Lupus erythematosus** | | |
| L93.0 Discoid lupus erythematosus | 20 | 91 |
| L93.1 Subacute cutaneous lupus erythematosus | 2 | 6 |
| L93.2 Other local lupus erythematosus | 3 | 8 |
| **M32 Systemic lupus erythematosus** | | |
| M32.1 Systemic lupus erythematosus with organ or system involvement | 30 | 32 |
| M32.8 Other forms of systemic lupus erythematosus | 5 | 6 |
| M32.9 Systemic lupus erythematosus, unspecified | 83 | 280 |
| M32.90 Systemic lupus erythematosus, unspecified, Multiple sites | 1 | 3 |

**Count of self-reported autoimmune condition**

| Self-reported disorder | Number of people |
| --- | --- |
| Systemic lupus erythematosis/sle | 453 |

**Count of self-reported prescription medications**

| Disease | Medication | Count |
| --- | --- | --- |
| Systemic Lupus Erythematosus | prednisolone | 2,310 |
|  | methotrexate | 2,096 |
|  | hydroxychloroquine | 590 |
|  | mycophenolate | 174 |
|  | tacrolimus | 172 |
|  | prednisolone product | 139 |
|  | ciclosporin | 50 |
|  | mtx - methotrexate | 36 |
|  | methylprednisolone | 19 |
|  | azt - azathioprine | 13 |
|  | myfortic 180mg gastro-resistant tablet | 13 |
|  | tacrolimus monohydrate 0.03% ointment | 13 |
|  | ciclosporin product | 1 |
